# Influenza vaccination allocation in tropical settings under constrained resources

**DOI:** 10.1101/2024.02.08.24302551

**Authors:** Joseph L Servadio, Marc Choisy, Pham Quang Thai, Maciej F Boni

**Author notes:** corresponding author: Joseph L Servadio, Department of Biology, Temple University, Philadelphia, PA, United States.

## Abstract

Influenza virus seasonality, synchronicity, and vaccine supply differ substantially between temperate and tropical settings, and optimal vaccination strategy may differ on this basis. Most national vaccine recommendations focus on high-risk groups, elderly populations, and healthcare workers despite previous analyses demonstrating broad benefits to vaccinating younger high-contact age groups. Here, we parameterized an age-structured non-seasonal asynchronous epidemiological model of influenza virus transmission for a tropical low-income setting. We evaluated timing and age allocation of vaccines across vaccine supplies ranging from 10% to 90% using decade-based age groups. Year-round vaccination was beneficial when comparing to vaccination strategies focused on a particular time of year. When targeting a single age-group for vaccine prioritization, maximum vaccine allocation to the 10-19 high-contact age group minimized annual influenza mortality for all but one vaccine supply. When evaluating across all possible age allocations, optimal strategies always allocated a plurality of vaccines to school-age children (10-19). The converse however was not true as not all strategies allocating a plurality to children aged 10-19 minimized mortality. Allocating a high proportion of vaccine supply to the 10-19 age group is necessary but not sufficient to minimize annual mortality as distribution of remaining vaccine doses to other age groups also needs to be optimized. Strategies focusing on indirect benefits (vaccinating children) showed higher variance in mortality outcomes than strategies focusing on direct benefits (vaccinating the elderly). However, the indirect benefit approaches showed lower mean mortality and lower minimum mortality than vaccination focused on the elderly.

**Significance statement:** Influenza exhibits strong annual seasonality in temperate countries, but less consistent and predictable patterns in tropical countries. Many tropical countries are low-income countries with low influenza vaccine coverage. Globally, influenza vaccines are recommended for elderly adults and vulnerable groups, though evidence has shown that vaccinating school-age children is beneficial due to their high rates of social contact. Our modeling study evaluated whether age-based vaccine allocations can effectively minimize population influenza mortality in a tropical country with constrained resources and little seasonality. Prioritizing school-aged children for vaccination minimized mortality, with secondary emphasis on elderly adults. These benefits are most apparent under low vaccine supplies and can inform most effective ways to develop or expand influenza vaccination campaigns in low-income tropical settings.

## Introduction

Influenza remains a persistent public health challenge globally causing up to 650,000 deaths annually [1]. It is present worldwide and prior to the emergence of SARS-CoV-2 was the most deadly infectious disease in many high-income [2–4] and some lower-income countries [5]. Morbidity and mortality from influenza can be mitigated through regular administration of vaccines. In high-income countries (HICs), between 28% and 42% of residents are vaccinated for influenza annually [6–8], primarily targeting elderly adults and those with underlying health conditions [9] with the majority of vaccines given prior to the onset of the winter influenza season. Most lower and lower-middle income countries (LMICs) have influenza vaccine coverage below 10% [10] with age/group targeting similar to higher income countries and no specific annual timing for vaccination campaigns.

When introducing or increasing coverage of influenza vaccines in lower-income settings, three major differences in influenza epidemiology to consider between HICs and LMICs are that LMICs have younger populations, irregular and unpredictable seasonality of influenza (if they are tropical or sub-tropical), and lower vaccine supplies. Timing vaccination just prior to an epidemic is important due to evidence of waning of vaccine-induced protection over time [11,12], and optimizing age-specific allocation is critical for vaccine distribution as influenza transmission as well as morbidity and mortality vary with age. Countries with established influenza vaccination programs consistently encourage vaccination during a particular time of year and seek to optimize vaccine recommendations based on age, exposure, or vulnerability to maximize the vaccine rollout’s mortality reductions. This planning is of course predicated on sufficient supply and population willingness to be vaccinated.

Almost universally, emphasis is placed on vaccinating populations at highest risk of exposure or severe outcomes, including healthcare workers, young children, and elderly adults. This is seen across HICs and LMICs by promoting vaccination in particular high-risk groups as part of a general voluntary vaccination approach [13–18]. Prioritization based on advanced age or health status was likewise reflected in COVID-19 vaccination rollouts in 2021 [19,20]. However, compared to COVID-19, influenza has a shallower age-severity curve and vaccination approaches can be more flexibly designed to target different age groups based on both contact mixing and severity considerations. As an example, an alternate strategy that focuses influenza vaccination on school-age children – a group with high levels of in-group and out-group social contact – was followed in Japan in the 1970s and 1980s, with some evidence of success in reducing nationwide influenza mortality [21]. Additional research from the United Kingdom found it to be a cost-effective method of reducing influenza mortality [22]. Other studies aiming to compare age-based vaccination strategies for influenza have typically found that vaccinating school-age children is advantageous due to their large contribution to transmission [23–29]. This choice between focusing vaccination on high-risk groups versus high-contact groups is a classic question in vaccine allocation for respiratory pathogens and depends strongly on overall transmission rate, the age-structure of the population and social contacts, and the age-morbidity association [30].

In this study, we parameterized an age-structured mathematical model of influenza transmission to the asynchronous non-annual epidemiology of tropical influenza in Vietnam, where influenza vaccination is currently being planned. Vietnam’s high population, inconsistency of influenza epidemic timing, and presence of both subtropical and tropical climates make it a suitable case study where results may apply to other subtropical and tropical countries, particularly within southeast Asia. We evaluated vaccine allocations in this context to determine the optimal allocation of influenza vaccines to see if this differs from recommendations typical in temperate high-income settings.

## Results

The mathematical model developed in this study includes three (sub)types of influenza, decade-based age groups, and compartments for those who received an influenza vaccine (Supplemental text S1, Figure S1). Fifty parameterizations of this dynamic epidemiological model of tropical influenza transmission were generated that achieved (1) sustained co-circulation of three influenza (sub)types: influenza A/H1N1, influenza A/H3N2, and influenza B; (2) non-damped oscillation of all three subtypes; (3) no synchronicity or phase-locking between any two subtypes; (4) stationary annual all-influenza attack rates between 15% and 35%; and (5) no clear dominance of any one subtype (Supplemental text S1). These dynamics are based on a well-characterized tropical influenza time series from Vietnam (4641 incidence data points over ten years). Across the fifty parameter scenarios, the median annual attack rate is 22.6% (IQR: 21.6-23.8%), and median annual mortality is 15,499 deaths (IQR: 14,816-16,427) in a population of 100 million, with 46.6% (IQR: 46.2 – 47.1) of deaths occurring in the 70+ age group and 30.9% (IQR: 30.8 – 31.0) occurring in the 60-69 age group. Adjusting for population, this is consistent with a relatively severe influenza season in the United States. Figure 1 shows examples of the simulated influenza scenarios in which we evaluated vaccine timings and age allocations as well as distributions of parameters among these 50 sets.

**Figure 1.**
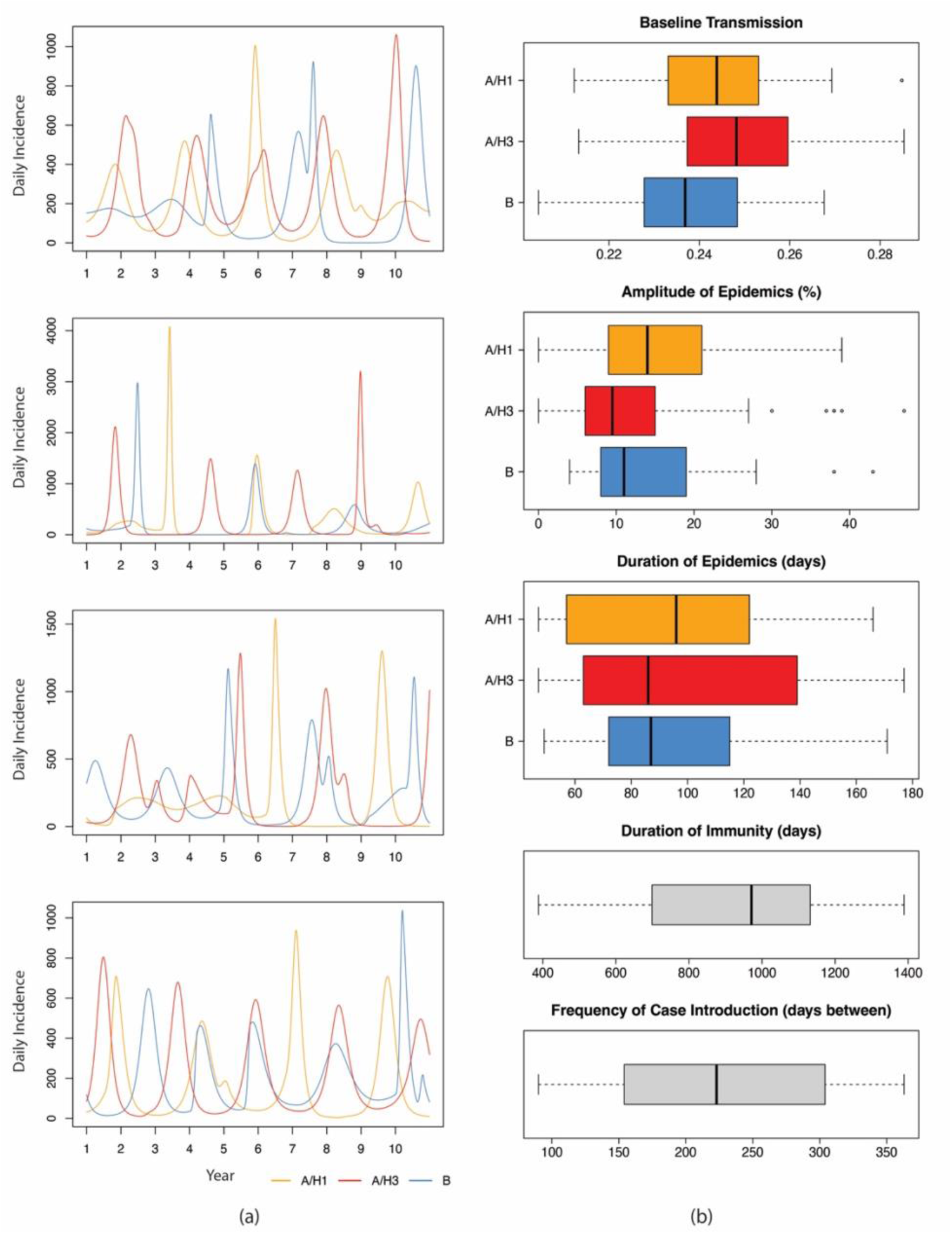
Model parameterizations epidemiologically consistent with observed influenza dynamics in Vietnam. (a) Four example model incidence trajectories, with daily incidence values for influenza A/H1N1, A/H3N2, and B over a ten-year period. The parameterizations leading to these trajectories were used in modeling to evaluate effectiveness of vaccination strategies. (b) Distributions of the parameters comprising these parameter sets.

Some evidence was found for a preferred time of year for vaccinating. Among vaccine programs spanning four months, changing the starting month reduced mortality by between 219 (1.6%) and 1,089 (32.4%) deaths per year across seven vaccine supplies.

Larger differences were seen under higher vaccine supplies. The starting month minimizing mortality was January for all except one vaccine supply (40%), where February was the starting month minimizing mortality (Figure S2). Implementing a year-round vaccination strategy, however, led to the lowest average annual mortality; across vaccine supplies, a year-round vaccine campaign led to 0 to 7% fewer deaths than the 4-month vaccination campaign with lowest mortality. In all evaluated scenarios, increasing vaccine supply proved substantially more influential in reducing influenza mortality compared to aiming to vaccinate during a particular time of year.

When considering a single age-group prioritization scheme, our analysis shows benefits of targeting school-age children for reducing influenza mortality among all age groups in a population. Across different vaccine supplies, we allocated vaccines to a single age group until full coverage was achieved; remaining vaccines were allocated proportionally across the other seven age groups. For eight of the nine vaccine supplies, lowest population mortality was achieved by prioritizing the 10-19 age group (Figure 2). Prioritizing the two oldest age groups (60-69 and 70+) typically also led to higher reductions in mortality compared to other age group prioritizations, and prioritizing the 70+ age group produced the lowest average annual mortality for a vaccine supply that covered 20% of the population. Differences in mortality based on the age group prioritized is more pronounced under lower vaccine supplies; compared to age-proportional vaccination, prioritizing the 10-19 age group leads to a reduction in mortality that ranges from 2.3% (20% supply) to 23.3% (60% supply). This is consistent with the well-known trade-off in respiratory disease vaccine allocation where the two major approaches to reduce disease burden are (1) reducing severity directly by vaccinating high-risk/vulnerable groups, and (2) reducing severity indirectly by reducing overall incidence which is achieved by vaccinating high-contact groups.

**Figure 2.**
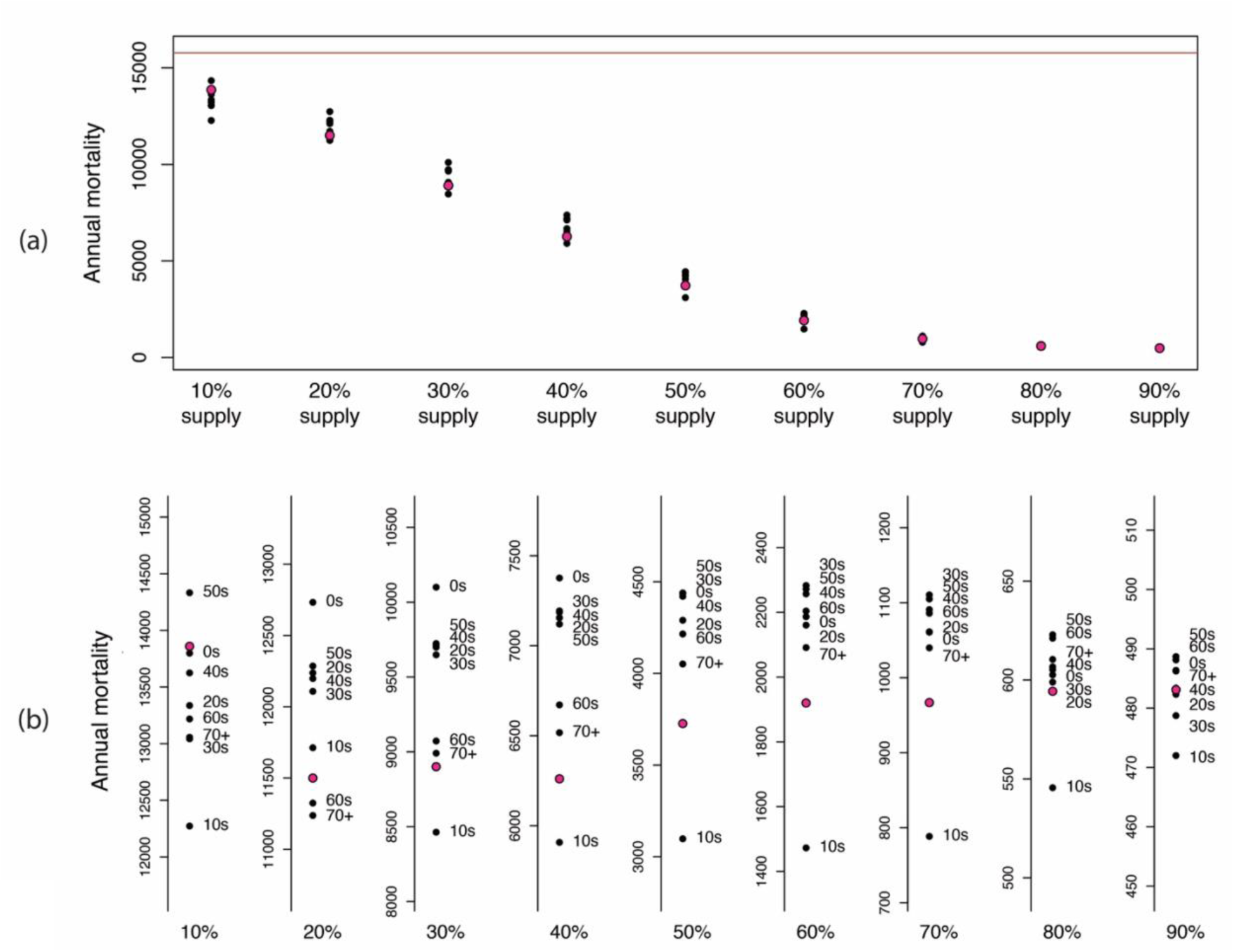
Expected annual mortality from prioritizing ten-year age bands (e.g. “20s” or “50s”) for vaccination. Under varying vaccine supplies, one age group was vaccinated until 100% coverage was reached, and remaining doses were distributed proportionally to the other seven age groups. Panel (a) shows eight black dots for the eight age-group prioritizations and one red dot for age-proportional distribution. The red line indicates mortality with no vaccination. Panel (b) shows zoomed-in and labeled mortality results for better distinction across age groups.

Even though single-age prioritization is a natural point of focus and often a default strategy considered in public health planning, allocating the vaccine supply to multiple key age groups may achieve further reductions in burden. We evaluated precise vaccine allocations across the eight age groups – 74,500 in total – chosen to approximate a brute-force approach to covering all possible age-group coverage combinations via a modified Latin Hypercube approach (see Methods and supplemental text S2). These evaluations show that a substantial lowering (up to a factor of 2.0) of mortality can occur when finding an optimal allocation for a given vaccine supply compared to a nonoptimal allocation. Figure 3 shows the distribution of annual mortality resulting from all tested allocations of vaccines for nine vaccine supplies. Greater benefits of age-specific allocation, when comparing to distributing vaccines proportionally across ages, are easily seen when vaccine supplies are low (≤50% supply). When comparing allocations that primarily vaccinate school-aged children (10-19) and the two oldest age groups (60+), primarily vaccinating younger children can lead to lower mortality, but with a wider range of possible mortality values (Figure 3b). This is seen further when examining the allocations that exclusively vaccinate either the two youngest or two oldest age groups for a vaccine supply available for 10% of the population (Figure 3c). Among the thirteen allocations exclusively vaccinating the oldest and youngest age groups, the lowest mortality is seen in one of the allocations exclusively vaccinating the two youngest age groups; however, all of the allocations exclusively vaccinating the oldest age groups lead to close-to-optimal low mortality due to the low variance of mortality outcomes when vaccination strategy aims for direct benefits over indirect benefits.

**Figure 3.**
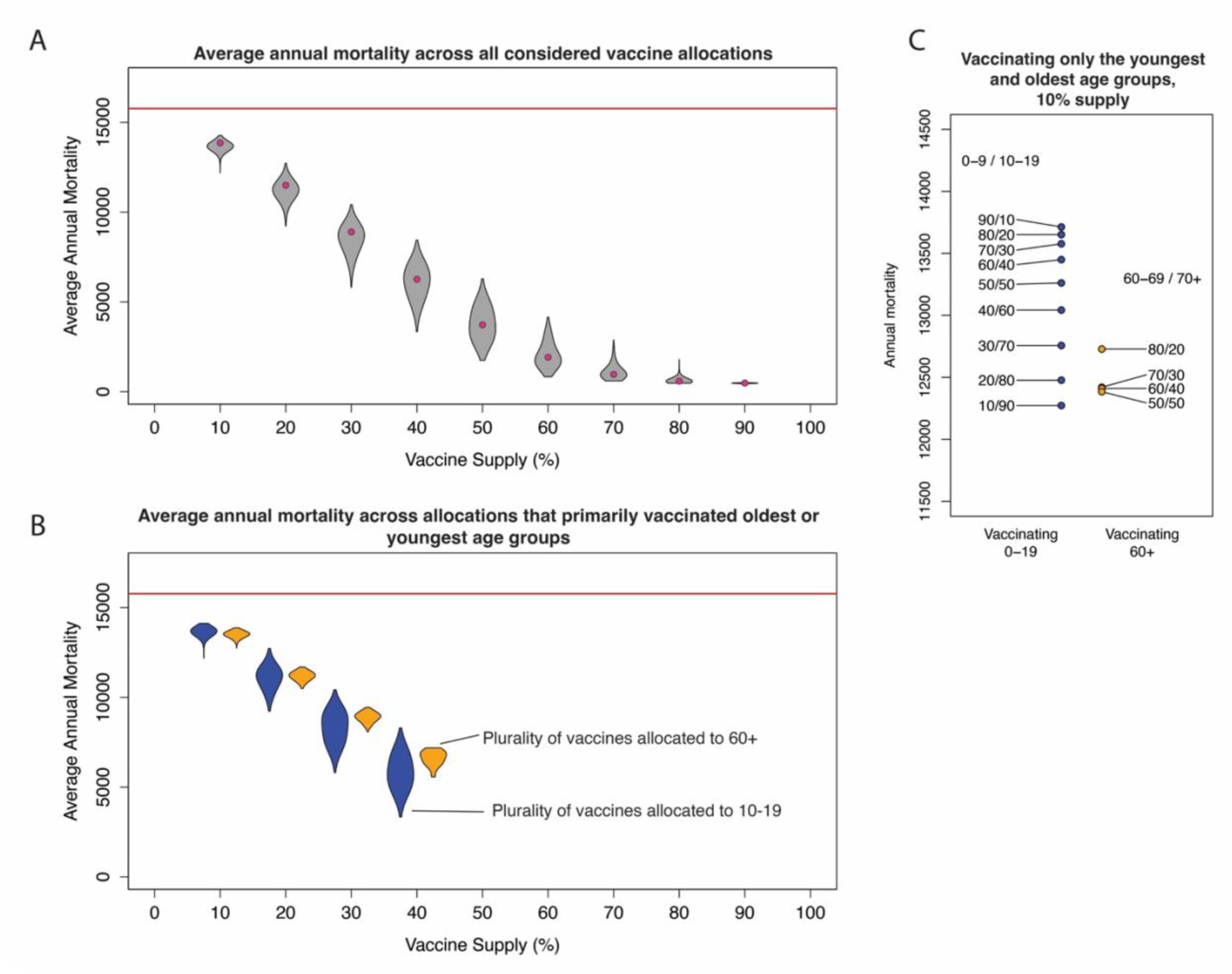
Mortality under various vaccine supplies and allocations. (a) Mortality among all considered vaccine allocations (N = 74500) for vaccine supplies for 10-90% of the population. Red points within the violin plots represent mortality if vaccines are allocated across age groups proportionally. (b) Mortality among the subsets of allocations represented in (a) that reserve the largest portion of vaccines for the 10-19 age group (blue) or two oldest age groups (orange). No vaccine allocations exist allocating a plurality of vaccines to the 60-69 or 70+ age groups when vaccine supply exceeds 40%. (c) Mortality under a vaccine supply for 10% of the population that exclusively vaccinates the two youngest age groups (left, blue) and two oldest age groups (right, orange). The percentage of vaccines allocated to the 0-9/10-19 age group as well as the percentage allocated to the 60-69/70+ age groups are indicated next to each point.

The vaccine allocations that best reduce mortality differ by vaccine supply, as shown in Figure 4. Under the lowest vaccine supplies, allocations that primarily focus on school-age children (10-19) were most effective, with some inclusion of elderly adults (70+) and adults of working age (20-39). These age groups represent those most likely to experience influenza mortality and those who have high degrees of social mixing within the population. Under more moderate vaccine supplies (available for 30-50% of the population), primarily vaccinating individuals between 10 and 49, largely representing school-age children and working adults, proved most effective. For vaccine allocations greater than 60%, vaccine allocations leading to the greatest reduction in mortality more closely resembled those proportional to the population age structure (Figure 4).

**Figure 4.**
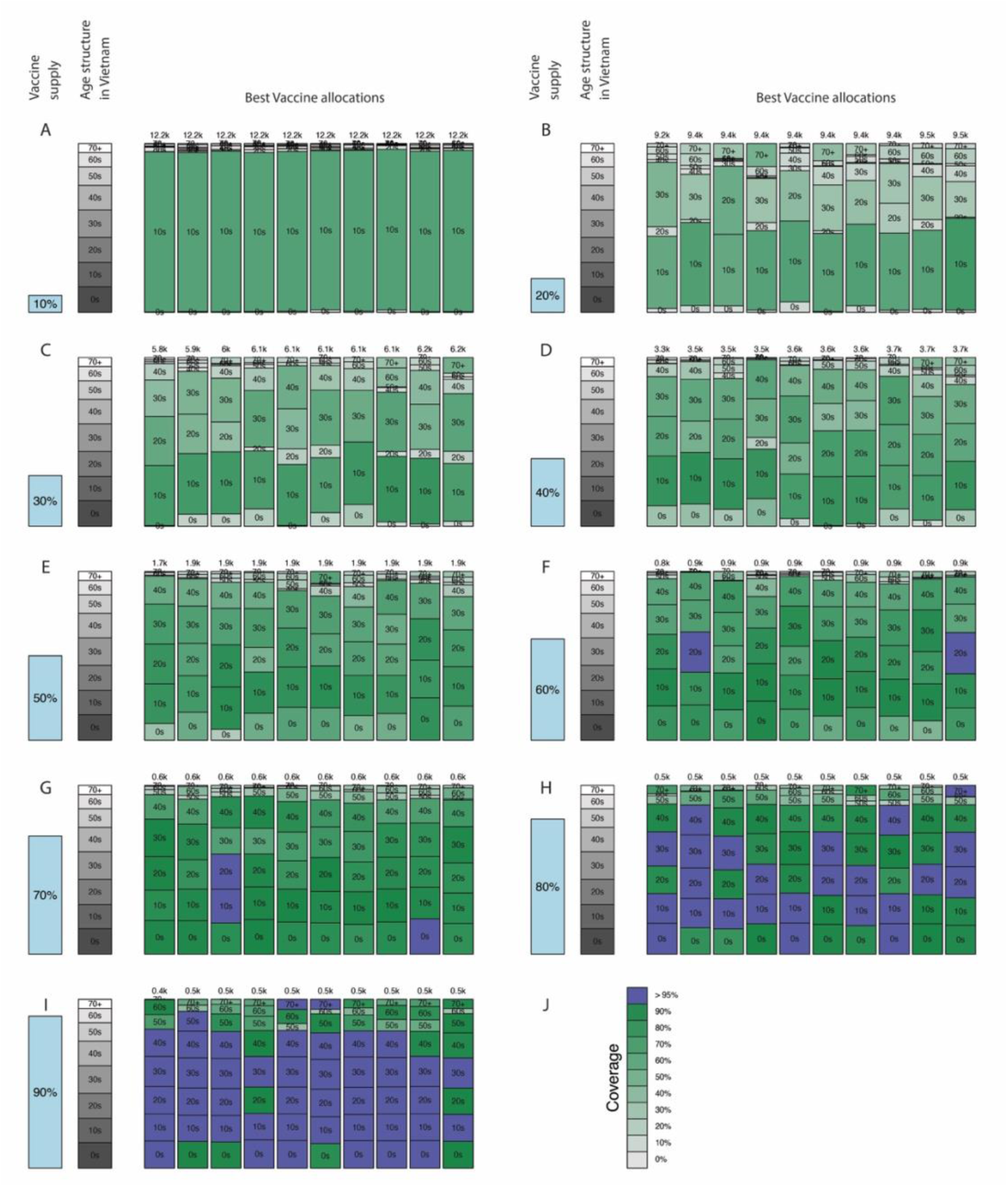
Optimal vaccine age-allocations across various vaccine supplies shown in panels A-I. In each panel, the light blue bar representing vaccine supply is shown next to the population age distribution, showing the relative sizes of the vaccine supply and population age groups. The ten blue/green bars on the right of each panel show the ten age allocations that minimize mortality, ranked from left to right by average annual total mortality. Annual mortality numbers for each allocation are shown at the top of each column. Blue age allocation boxes indicate vaccine coverage above 95% for that age group; age group coverage for green boxes corresponds to the color bar in panel J.

Prioritizing school-age children is necessary but not sufficient for minimizing mortality, particularly at low vaccine supplies. Among the vaccine allocations that prioritize school-age children, the vaccine distribution among the remaining doses across the other seven age groups can lead to notable differences in mortality. There are no straightforward monotonic relationships between age-group vaccine allocation and population mortality as can be seen clearly in Figure 5. Allocations achieving high vaccine coverage in the 10-19 age group need to also distribute adequate doses to adults 60+ to minimize mortality. This new finding on sufficiency of a certain age-group focus in vaccination programs – along with the wider variance in mortality when prioritizing school aged children – shows the importance of proper planning for allocating the entire vaccine supply.

**Figure 5.**
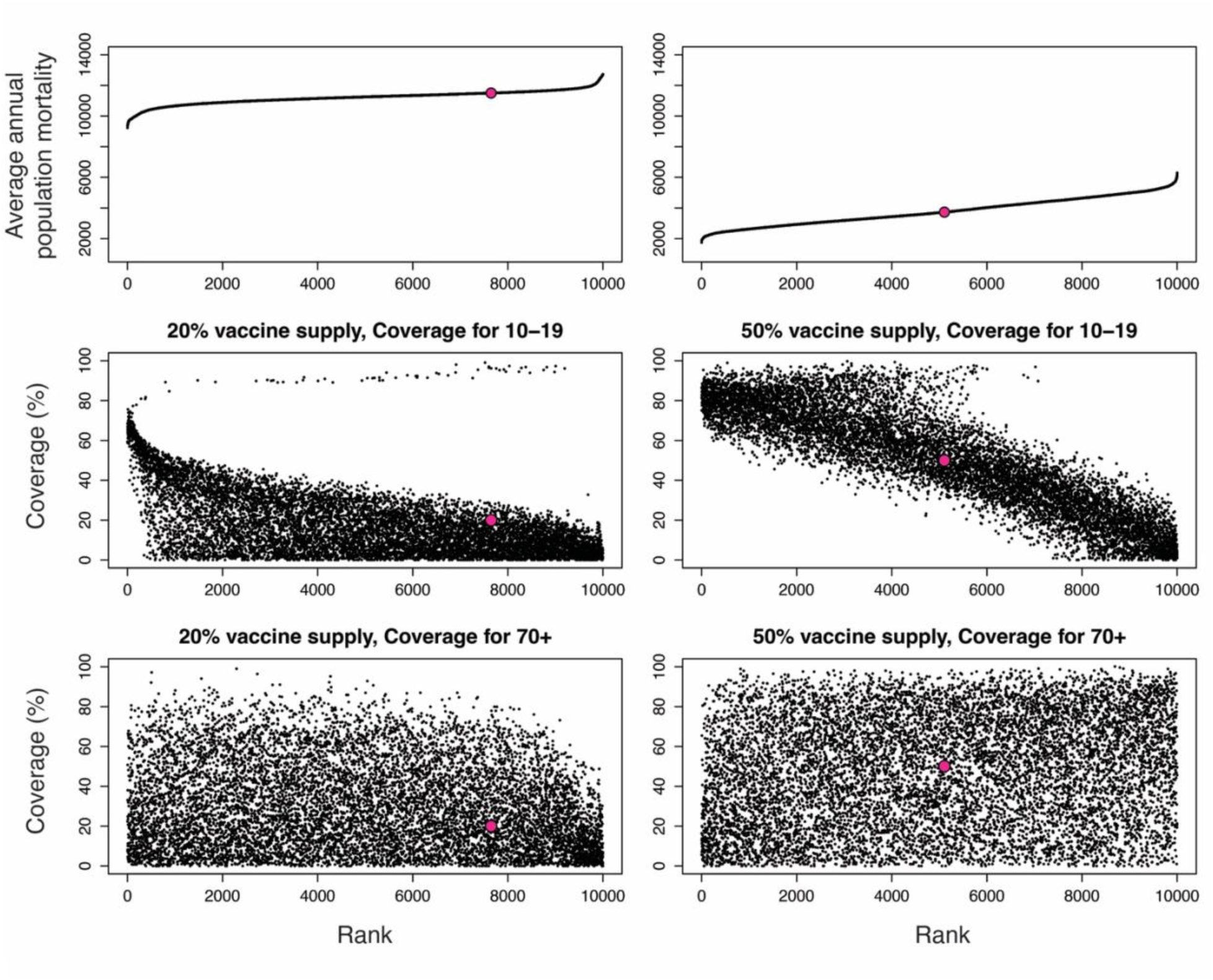
Vaccination coverages in two age groups (10-19, 70+) across all considered allocations of two vaccine supplies (20%, 50%), ranked by average annual population mortality. Red dots in each panel indicate the allocations with proportional vaccination across all age groups. Top row: Average annual mortality for each allocation. Middle row: Vaccination coverages for the 10-19 age group among the ranked allocations. The general downward slope shows the importance of vaccinating this age group for reducing population mortality, but the low rank of many allocations with high coverage for this age group with 20% supply (left) show that saturating this age group and leaving others unvaccinated is not guaranteed to be optimal. Bottom row: Vaccination coverages for the 70+ age group among ranked allocations. Full results, across all vaccine supplies and all age groups, are shown in Figure S3.

Our analyses assumed the effectiveness of influenza vaccines and duration of vaccine-induced immunity consistent with previous literature [31–36]. We repeated our analyses varying each of these to determine whether the vaccine allocations minimizing mortality change with different effectiveness values or immune durations (full details in Supplemental text S3). If vaccine effectiveness is reduced from 50% to 40% or 30%, targeting elderly adults typically led to lowest annual mortality values, particularly when vaccine supplies are lower. This suggests a new efficacy-dependent allocation that would need to be validated: majority allocation to older age groups during strain-mismatch years and majority allocation to younger age groups during other years. Under increased vaccine effectiveness, prioritizing school-age children remained advantageous for minimizing mortality (Figures S4 and S5). Similarly, reducing the assumed 9-month average duration of immunity to 6 to 7 months shifted the optimum vaccination strategy to one that prioritizes elderly adults while increasing immune duration in the model kept school-age children as the key group for prioritization (Figures S6 and S7). We also examined changes in age structure, comparing the age structures of Vietnam, the United States, and intermediate age structures. The benefits of primarily vaccinating the 10-19 age group, particularly at low vaccine supplies, were seen across all five age structures (Figures S8, S9), indicating that age-structure differences in HICs and LMICs should not have a substantial effect on the choice of optimal vaccine policy.

## Discussion

This study aimed to identify vaccine distribution strategies that most effectively reduce annual mortality in the absence of annual seasonality. This absence of seasonality is most typically seen in tropical countries, many of which are LMICs with currently low vaccine coverage. We used influenza dynamics from Vietnam to inform a non-seasonal asynchronous influenza model and evaluated various vaccine allocations under different supply constraints, resembling the development or expansion of a national vaccine program. A clear time of year was not identified that would most effectively reduce influenza mortality, and age-structure difference between tropical and temperate settings did not seem to influence vaccine policy choice. Age-based allocations that prioritize school-aged children were most effective at reducing overall mortality, and benefits of vaccinating working-age adults and elderly adults were also seen (Figure 4). The benefits of prioritizing particular age groups are most prominent when vaccine supplies are low, emphasizing their importance when first establishing a vaccine program. Under higher vaccine supply, allocating vaccines proportionally across age groups is an adequate burden reduction approach. Our finding of the advantages in prioritizing school-age children differs from the typical recommendations that prioritize elderly adults but is consistent with previous studies [24,25,37].

Strategies that led to the greatest reduction in mortality included vaccinating school-aged children and elderly adults, the groups with the highest social mixing in the population [38,39] and mortality risk [40,41], respectively. Children have also previously been found to have higher observed influenza incidence [42]. The benefits of prioritizing these two groups rather than allocating vaccines proportionally are greatest when considering the lowest vaccine supply (available for 10% of the population). Vaccinating working-age adults, particularly those aged 30-39, was also beneficial under lower vaccine supplies, which is supported by previous literature [24]. The benefits of prioritizing school-aged children depended on factors such as vaccine supply, vaccine effectiveness, and average duration of vaccine-inferred immunity. Decreasing vaccine effectiveness or duration of vaccine-induced immunity, led to strategies that favor primarily vaccinating elderly adults being the most effective (Figures S4-S7). Because vaccine effectiveness can fluctuate across years, it is possible that the optimal age-based strategies will vary across years.

The World Health Organization’s position on influenza vaccination emphasizes annual vaccination for elderly adults because they are at the highest risk of death [43,44]. Some previous studies regarding vaccine strategies have also found that prioritizing elderly adults is most beneficial for both influenza [45] and COVID-19 [46]. Other studies, however, have considered the benefits of prioritizing school-age children based on their high level of social mixing both within and outside of their age group [47]. A study from the United Kingdom found vaccinating children to be of greater overall benefit, with reallocating vaccines to elderly adults being less cost effective [22]. The decision in practice to not focus vaccination on school-age children may result from increased barriers to uptake such as inconvenience, complacency, unawareness of the magnitude of benefit, or perceptions of influenza-associated risk [48,49].

Vaccine supply – a variable that is not considered in optimal allocation problems in wealthier countries – plays a critical role in determining optimal allocation in a resource-constrained setting. We focused on supply to consider the development of a vaccine program in a tropical country that currently does not currently achieve high vaccination coverage, such as Vietnam. Most previous studies examining effective ways to vaccinate a population for influenza or other respiratory viruses primarily investigate population coverage [50,51] or distribution of a fixed supply [25,52], with some giving attention to vaccine supply [24,46,53]. By considering how to allocate among nine vaccine supplies, our results are more applicable to policy makers and health professionals, showing how optimal allocations can differ across supply levels (Figure 4). We assume perfect adherence to each allocation. In practice however, some allocations may be less feasible due to age-differentiated willingness to be vaccinated or access to vaccination (such as ease of vaccinating in a school setting compared to visiting a clinic for vaccination).

In the context of an existing, but limited, vaccine program, an open question in optimal vaccine allocations is how to most effectively use an expanding vaccine supply. By examining a broad range of vaccine supplies, we showed how optimal allocations change when establishing a small vaccine program and then when expanding a current program. When developing a program in a tropical country, our results support primarily allocating a very limited supply to school-age children and elderly adults. Under more moderate supplies, it is beneficial to include working-age adults. At the highest supply levels, it is sufficient to vaccinate proportionally across age groups, with less benefits from opting for an age-based allocation.

Our model accounts for age-based influenza dynamics but does not consider differences across locations. These can include asynchronous influenza epidemics among northern, central, and southern Vietnam [54] or differences in vaccine coverage between urban and rural communities [55]. Our model also only considers age for both hospitalization and mortality risk as well as for vaccine priorities. Differences in severity have been seen based on sex [56], and other groups that may benefit from vaccine prioritization include healthcare workers (recommended in Vietnam [57]) and people with underlying health conditions [58]. Additional simplifying assumptions were made regarding vaccine-induced immunity. By having a single set of vaccine compartments, we assumed that all protection is lost after vaccine immunity wanes, though immunity may wane only partially over time [59,60]. We also assumed uniform vaccine effectiveness for all age groups. There is evidence of lower vaccine effectiveness among elderly populations [61], though this finding is not universal [62]; therefore, our analysis may be overestimating the benefits of vaccinating to elderly adults.

Few models exist for tropical influenza circulation with coexistence of distinct types and subtypes [63]. As a result, we used a model with 50 parameterizations that reflect influenza dynamics in Vietnam and can account for uncertainty in some parameters [25]. In our analysis, prioritizing school-age children for annual influenza vaccination leads to the greatest reductions in annual mortality in the implementation of a newly expanded vaccine program. Characteristics of the vaccine, notably its average duration of inferred immunity and efficacy, may impact how to allocate vaccines in a way that minimizes mortality, particularly under low vaccine supplies. These findings are likely to be of interest to many tropical countries that experience irregular timing of influenza epidemics and with plans to increase currently low influenza vaccine coverage.

## Methods and Materials

This study focuses on vaccine strategies in Vietnam, a LMIC with a population of approximately 100 million. Vietnam has a subtropical climate in the northern half of the country and a tropical climate in the southern half. Previous research has not identified strong predictable cycles in influenza incidence for any (sub)type in any of northern, central, or southern Vietnam [54]. Vietnam currently has low influenza vaccine coverage, but there is interest in expanding coverage.

### Mathematical model

Since only one known model exists that fits three co-circulating (sub)types of influenza in a tropical region [63], we adapted our previous mathematical model, which was fit for a single nonannual pathogen [54], to match the circulation patterns of multiple types and subtypes of influenza, using fifty separate parameterizations for robustness. In our adapted model, each (sub)type follows SIRS dynamics with a two-stage infected class and a four-stage recovered class [64]. (Figure S1, text S1). Periods of increased transmission occur irregularly, with intervals between periods of increased transmission drawn stochastically from a previously fit normal distribution of epidemic timings from ten years of sentinel surveillance from fifteen hospitals located throughout Vietnam [54]. We incorporated previously estimated levels of cross-immunity among (sub)types [63]. Single cases are moved from the Susceptible class to an Infected class at regular intervals to represent case importation. Hospitalization can occur during infection, using a hospitalization fraction based on data from the US Centers for Disease Control and Prevention (CDC) [65]. Death occurs from the hospital at a rate based on CDC data [65].

We divided the population into eight decade-based age groups, with the last group consisting of adults 70 years and older, using published age demographics in Vietnam [66]. We incorporated a crude birth rate as reported by the World Bank [67] and an age-based natural death rate as reported by the World Health Organization [68]. Age-based contact mixing was incorporated in the model by using previously estimated age-based contact patterns for Vietnam [69,70].

To ensure that our model allows all three (sub)types of influenza to co-circulate with asynchronous behavior, we parameterized the model using known epidemiological quantities in Vietnam such as annual attack rate and variance in timings between epidemic peaks. Using a standard parameter-space search approach, we drew sets of parameters from uniform distributions and retained 50 sets that produced model dynamics resembling those seen in Vietnam, allowing asynchronous nonannual co-circulation of the three (sub)types. Parameter values from literature are listed in Table S1 and full details of the parameter-space search can be found in Supplemental text S1.

### Vaccine implementation

Our model allows vaccination from the Susceptible and Recovered compartments (Figure S1). We applied vaccine strategies to the specified model, running the model for a 10-year period and using average annual mortality over the ten years to compare effectiveness of each. We organized our analyses around the central variable of vaccine supply and how best to allocate vaccines given supply constraints, considering supply relative to the population. A 10% vaccine supply, therefore, is interpreted as having enough doses to vaccinate 10% of the population. We considered vaccine supplies that cover 10% through 90% of the population. The first vaccine strategies investigated whether administering vaccines during a particular time of year would lead to the greatest reduction in average annual mortality.

In these scenarios, vaccines were assumed to be distributed proportionally across all age groups within the population. We considered each calendar month as a starting point for vaccine administration, and we considered vaccine administration durations of one through four months. Additionally, we considered a scenario where vaccines are evenly distributed to the population throughout the entire year.

The second set of vaccination scenarios considered prioritizing single age groups. For each of the nine specified vaccine supplies, we considered prioritizing each age group by vaccinating all members of that age group until 100% coverage is reached. The remaining vaccines were then distributed to the remaining seven age groups proportionally to their respective sizes.

The final set of vaccination scenarios considered ways to allocate all available doses across all eight age groups rather than prioritizing one age group. In other words, we performed – as completely as possible give computational limitations – a brute-force search over all possible age-allocations. The purpose of this was to find previously unexamined optimal ways to distribute or allocate available vaccines to the eight age groups. Full details of developing these allocations can be found in supplemental text S2. Briefly, for each vaccine supply, we sampled proportions (in [0,1]) in each age group for an assigned vaccine coverage and applied them to our model to evaluate mortality. We drew allocations, in the form of sets of eight coverages, using Latin hypercube sampling, with (1) a rejection mechanism for allocations that require major rescaling, and (2) an added constraint to ensure that >95% coverages for individual age groups were achieved in the sampling. We drew 10,000 allocations for supplies ranging between 10 and 60 percent of the population and then 7500, 5000, and 2000 allocations for 70, 80, and 90 percent supplies, respectively, since fewer feasible allocations exist at those high coverages. Each allocation was evaluated for each of the 50 epidemiologically consistent parameter sets for Vietnam, and we averaged the results across parameter sets for each allocation. We also considered an allocation where doses are distributed proportionally to the relative sizes of age groups to compare specific age-based vaccine allocations to naively distributing vaccines uniformly throughout the entire population.

All analyses were run in R version 4.2.1 [71]. We used the ‘Rcpp’ package [72] for coding and implementing the model, the ‘lhs’ package [73] for Latin Hypercube Sampling, and the ‘parallel’ package for computational efficiency.

## Data Availability

Data generated in this study and relevant code are available at github.com/jlservadio/VN_Influenza_Vaccination

