## Supplemental Text for "Influenza vaccination allocation in tropical settings under constrained resources"

### Text S1. Model specification

Few models exist that attempt to fit three co-circulating (sub)types of influenza in a tropical region [1], so we adapted our previous model fit for a single nonannual pathogen [2] to fit multiple types and subtypes of influenza. In our adapted model, each (sub)type follows SIRS dynamics with a two-stage infected class and a four-stage recovered class to allow duration of infection and duration of infection-induced immunity to be Erlang distributed [3]. (Figure S1). Periods of increased transmission occur irregularly, with intervals between periods of increased transmission drawn stochastically from a previously fit normal distribution of epidemic timings from ten years of sentinel surveillance from fifteen hospitals located throughout Vietnam [2]. Infection with one (sub)type confers partial immunity to the others, using quantities of partial immunity estimated in previously published literature [1]. Single cases are sporadically introduced to the population at fixed intervals to represent case importation by moving an individual from the Susceptible compartment to an Infected compartment, alternating across (sub)types and age groups. Hospitalization can occur following the first infection stage, using a hospitalization fraction based on data from the US Centers for Disease Control and Prevention (CDC) [4]. Death occurs from the hospital at a rate based on CDC data [4]; we assumed that deaths only occur after hospitalization.

We incorporated age structure in the model by dividing the population into eight decade-based age groups, with the last group consisting of adults 70 years and older, using published age demographics in Vietnam [5]. We incorporated a crude birth rate as reported by the World Bank [6] and an age-based natural death rate as reported by the World Health Organization [7]. Risk of hospitalization and death vary by age group based on reported hospitalization and death fractions reported by the CDC [4]. Age-based contact mixing was incorporated in the model by using previously estimated age-based contact patterns for Vietnam [8,9].

The model is specified by the following equations, with parameters used shown in Table S1:

$$\frac{dS_{\tilde{a}}}{dt} = -\sum_{a}\sum_{s}\eta_{a\tilde{a}}\beta_s(t)S_{\tilde{a}}(I_{as}^1 + I_{as}^2 + vI_{as}^1 + vI_{as}^2) + \sum_{s}\frac{\rho}{4}R_{\tilde{a}s}^4 + \sum_{s}\frac{\rho}{4}vR_{\tilde{a}s}^4 + \rho_v V_{\tilde{a}}$$

$$\frac{dI_{\tilde{a}\tilde{s}}^1}{dt} = \sum_{a}\eta_{a\tilde{a}}\beta_{\tilde{s}}(t)S_{\tilde{a}}\,(I_{a\tilde{s}}^1 + I_{a\tilde{s}}^2 + vI_{a\tilde{s}}^1 + vI_{a\tilde{s}}^2)$$

$$+ \sum_{a}\sum_{s\neq\tilde{s}}\sum_{i=1}^{4}\eta_{\tilde{a}a}\beta_{\tilde{s}}(t)R_{as}^i(I_{a\tilde{s}}^1 + I_{a\tilde{s}}^2 + vI_{a\tilde{s}}^1 + vI_{a\tilde{s}}^2) - \frac{hv}{2}I_{\tilde{a}\tilde{s}}^1 - \frac{(1-h)v}{2}I_{\tilde{a}\tilde{s}}^1$$

$$\frac{dI_{\tilde{a}\tilde{s}}^2}{dt} = \frac{(1-h)v}{2}I_{\tilde{a}\tilde{s}}^1 - \frac{v}{2}I_{\tilde{a}\tilde{s}}^2$$

$$\frac{dH_{\tilde{a}\tilde{s}}}{dt} = \frac{hv}{2}I_{\tilde{a}\tilde{s}}^1 - \frac{dv}{2}H_{\tilde{a}\tilde{s}} - \frac{(1-d)v}{2}H_{\tilde{a}\tilde{s}}$$

$$\frac{dR_{\tilde{a}\tilde{s}}^1}{dt} = \frac{v}{2}I_{\tilde{a}\tilde{s}}^2 + \frac{(1-d)v}{2}H_{\tilde{a}\tilde{s}} - \frac{\rho}{4}R_{\tilde{a}\tilde{s}}^1 - \sum_{a}\sum_{s\neq\tilde{s}}\eta_{a\tilde{a}}\beta_s(t)R_{\tilde{a}\tilde{s}}^1\,(I_{as}^1 + I_{as}^2 + vI_{as}^1 + vI_{as}^2)$$

$$\frac{dR_{\tilde{a}\tilde{s}}^i}{dt} = \frac{\rho}{4}R_{\tilde{a}\tilde{s}}^{i-1} - \frac{\rho}{4}R_{\tilde{a}\tilde{s}}^i - \sum_{a}\sum_{s\neq\tilde{s}}\eta_{a\tilde{a}}\beta_s(t)R_{\tilde{a}\tilde{s}}^i\,(I_{as}^1 + I_{as}^2 + vI_{as}^1 + vI_{as}^2)\,, i \in \{2, 3, 4\}$$

$$\frac{dV_{\tilde{a}}}{dt} = -\sum_{a}\sum_{s}\eta_{a\tilde{a}}\xi\beta_s(t)V_{\tilde{a}}(I_{as}^1 + I_{as}^2 + vI_{as}^1 + vI_{as}^2) - \rho_v V_{\tilde{a}}$$

$$\frac{dvI_{\tilde{a}\tilde{s}}^1}{dt} = \sum_{a}\eta_{a\tilde{a}}\xi\beta_{\tilde{s}}(t)V_{\tilde{a}}\,(I_{a\tilde{s}}^1 + I_{a\tilde{s}}^2 + vI_{a\tilde{s}}^1 + vI_{a\tilde{s}}^2)$$

$$+ \sum_{a}\sum_{s\neq\tilde{s}}\sum_{i=1}^{4}\eta_{\tilde{a}a}\xi\beta_{\tilde{s}}(t)vR_{as}^i(I_{a\tilde{s}}^1 + I_{a\tilde{s}}^2 + vI_{a\tilde{s}}^1 + vI_{a\tilde{s}}^2) - \frac{\xi hv}{2}vI_{\tilde{a}\tilde{s}}^1$$

$$- \frac{(1-\xi h)v}{2}vI_{\tilde{a}\tilde{s}}^1$$

$$\frac{dvI_{\tilde{a}\tilde{s}}^2}{dt} = \frac{(1-\xi h)v}{2}vI_{\tilde{a}\tilde{s}}^1 - \frac{v}{2}vI_{\tilde{a}\tilde{s}}^2$$

$$\frac{dvH_{\tilde{a}\tilde{s}}}{dt} = \frac{\xi hv}{2}vI_{\tilde{a}\tilde{s}}^1 - \frac{\xi dv}{2}vH_{\tilde{a}\tilde{s}} - \frac{(1-\xi d)v}{2}vH_{\tilde{a}\tilde{s}}$$

$$\frac{dvR_{\tilde{a}\tilde{s}}^1}{dt} = \frac{v}{2}vI_{\tilde{a}\tilde{s}}^2 + \frac{(1-\xi d)v}{2}vH_{\tilde{a}\tilde{s}} - \frac{\rho}{4}vR_{\tilde{a}\tilde{s}}^1$$

$$- \sum_{a}\sum_{s\neq\tilde{s}}\eta_{a\tilde{a}}\xi\beta_s(t)vR_{\tilde{a}\tilde{s}}^1\,(I_{as}^1 + I_{as}^2 + vI_{as}^1 + vI_{as}^2)$$

$$\frac{dvR^{i}_{\tilde{a}\tilde{s}}}{dt} = \frac{\rho}{4} vR^{i-1}_{\tilde{a}\tilde{s}} - \frac{\rho}{4} vR^{i}_{\tilde{a}\tilde{s}} - \sum_{a} \sum_{s \neq \tilde{s}} \eta_{a\tilde{a}} \xi \beta_s(t) vR^{i}_{\tilde{a}\tilde{s}} \left(I^1_{as} + I^2_{as} + vI^1_{as} + vI^2_{as}\right), i \in \{2, 3, 4\}$$

$$\beta_{\tilde{s}}(t) = \begin{cases} \beta_{0\tilde{s}} \left[1 + a_{\tilde{s}} \cos^{+}\left(\frac{2\,\pi\,(\varphi_{k\tilde{s}} - t)}{2\,\delta_{\tilde{s}}}\right)\right], \varphi_{k\tilde{s}} - \frac{\delta_{\tilde{s}}}{2} \leq t \leq \varphi_{k\tilde{s}} + \frac{\delta_{\tilde{s}}}{2} \\ \beta_{0\tilde{s}}, otherwise \end{cases}$$

$$\varphi_{k+1,\tilde{s}} = \varphi_{k\tilde{s}} + \tau_{k\tilde{s}}$$

where $\tilde{a}$ denotes any particular age group and $\tilde{s}$ denotes any particular (sub)type of influenza.

Expanding our model from one pathogen to three does not guarantee that all three influenza (sub)types co-circulate with asynchronous epidemic behavior (i.e. without reaching an endemic state). Influenza dynamics in Vietnam have showed subtypes A/H1, A/H3, and type B experience epidemic peaks of varying sizes that occur asynchronously (with B epidemics showing periods of flat incidence approximately half of the time) [2,10]. Therefore, we aimed to adapt our model to emulate asynchronous behavior to better reflect influenza dynamics in Vietnam. In doing so, we used the known annual attack rate [11] and previously inferred variances in timing of epidemic peaks to identify this model behavior. Because neither strong annual nor nonannual cycles were detected in Vietnam previously, we did not fit the model to the observed data since it is unrealistic to assume that the observed trends over ten years will be prescriptive or representative of future dynamics.

We used known epidemiological quantities in Vietnam (annual attack rate and variance in timings between epidemic peaks) to inform model parameterization. To account for the uncertainty in model parameters and to avoid producing results that are sensitive to assumed parameter values [12], we considered multiple parameterizations. We assumed a five-day duration of infection, and cross-immunity among subtypes based on previous literature [1]. For remaining model parameters (baseline transmission for each (sub)type $\beta_{0H1}, \beta_{0H3}, \beta_{0B}$; duration of infection-induced immunity $\frac{1}{\rho}$; magnitude $a$, timing $\tau_k$, and duration $\delta$ of epidemic peaks for each (sub)type; and frequency of sporadic case introduction), we conducted a random parameter search to identify parameter combinations that are epidemiologically consistent with influenza dynamics in Vietnam. The parameter search is in the form of an Approximate Bayesian Computation where a set of seven conditions must be satisfied for acceptance. We ran the model for a 20-year burn-in period, followed by a 10-year period where we compared model output to expected dynamics seen in Vietnam to identify realistic parameter sets. We classified parameter combinations as epidemiologically consistent with influenza dynamics observed in Vietnam if they produce stable forward dynamics with asynchronous co-circulation of the three (sub)types without settling into endemic equilibrium. In order to identify parameter sets meeting these conditions, we set seven criteria that must all be met:

i) Standard deviation of daily incidence values for each (sub)type are greater than 100 (in a population of one million)
ii) Pairwise correlations between pairs of (sub)types are less than 0.7 in absolute value
iii) Among the maximum daily incidence values of each (sub)type, the lowest maximum value is no less than 70% of the highest maximum incidence value.
iv) The three maximum daily incidence values must span at least one year (i.e. all three peaks cannot be in the same 365-day period)
v) Within each (sub)type, between 25 and 75% of daily incidence values are less than 100 (in a population of one million)
vi) All-influenza annual attack rates (combining all (sub)types) are between 15 and 35%.
vii) All-influenza annual attack rates are not monotonically increasing or decreasing over time.

Model parameters are listed in Table S1, including distributions from which we drew parameter values in the parameter search. We chose the first 50 parameter sets that satisfied all seven criteria described previously to assure our results are robust to realistic parameterizations.

We introduced vaccination into the model by creating a set of compartments for vaccinated individuals. In order to account for vaccines not only preventing infection but also reducing risk of severe outcomes (hospitalization or death) following infection, this included a set of infected, hospitalized, and recovered classes among those vaccinated (Figure S1). Based on previous research regarding vaccine protection against infection [13], hospitalization [14–16], and death [17], we assumed a 50% effectiveness in preventing infection, 50% effectiveness in preventing hospitalization from infection, and 50% effectiveness in preventing death from hospitalization. We also assumed an exponentially distributed duration of vaccine-induced immunity, with a mean duration of 270 days. Under this assumption, approximately half of vaccinated individuals retain protection after six months, and approximately a quarter retain protection after one year, which is broadly consistent with previous findings [18].

**Text S2. Assigning vaccine allocations**

In order to identify vaccine allocations that best reduce mortality, we examined a large number of allocations for each vaccine supply to see which allocation led to lowest average annual mortality. Each of these allocations consists of a set of eight numbers corresponding to the proportion of vaccines allocated to each age group, which can be algebraically translated into vaccine coverage proportions for each age group. Each set of eight numbers was randomly drawn to represent a wide variety of ways to distribute vaccines to eight different age groups. In order to assure that high vaccine coverage was considered for all age groups for each vaccine supply, we used a three-step process for each vaccine supply:

1. Draw Latin hypercube samples representing the percentage of each age group that is covered in vaccination (sets of eight numbers ranging between zero and one). Keep only those that yield vaccine totals above the specified amount but less than the specified amount plus two percentage points. Scale all eight values down to achieve the exact vaccine supply.
2. For each age group, repeat step 1 with the additional constraint of keeping only samples where coverage in that age group exceeds 95% (or 95% of the maximum possible coverage for the 10% vaccine supply). Repeat until each age group contains a minimum of 25 samples with high coverage.
3. Repeat step 1 until the desired total number of allocations is achieved.

We drew 10,000 total samples for supply levels ranging between 10 percent and 60 percent of the population. At higher supply levels, there are fewer possible feasible allocations, so fewer sampled allocations were needed to achieve a full range of possible coverage values. We drew 7,500 samples for the 70 percent supply, 5,000 samples for the 80 percent supply, and 2,000 samples for the 90 percent supply. We applied each allocation to the 50 parameter sets and averaged their average annual mortality to evaluate the effectiveness of each allocation in reducing mortality.

**Text S3. Sensitivity analyses**

We assumed a vaccine effectiveness of 50% against infection, against hospitalization if infected, and against death if hospitalized. While this is consistent with previous estimates for vaccine effectiveness in reducing infection and death, we examined different vaccine effectiveness values since it has been observed to change across years. To see if optimal strategies differ, we repeated our analyses using 30, 40, 50, 60, and 70% vaccine effectiveness. Under a vaccine effectiveness of 30% or 40%, targeting elderly adults generally led to lowest annual mortality, particularly when vaccine supplies were lower. As vaccine effectiveness increased to 50% or greater, the allocations leading to lowest mortality favored vaccinating younger age groups (Figure S4, S5). The magnitude of differences in vaccine allocations can be large; when examining the ten allocations with lowest mortality under a 10% supply, we see very few vaccines are allocated to the 10-19 group under 30% effectiveness, a mix of allocations with very low and very high doses allocated to the 10-19 group under 40% effectiveness, and nearly all vaccines allocated to the 10-19 group when effectiveness is 50% or higher (Figure S4).

Figure S5 shows the share of each age group that comprises the 10 allocations leading to lowest mortality across vaccine supplies and vaccine effectiveness values by averaging the percentage of doses allocated to each age group among the ten. Within each vaccine supply, increases in the share of doses allocated to younger age groups generally increases as vaccine effectiveness increases. This change is most substantial when vaccine supplies are lower since most effective allocations more closely resemble population demographics when vaccine supplies are high.

Estimates regarding vaccine immunity waning are also uncertain. Previous studies have produced varying estimates for the duration of immunity following vaccination. Therefore, in addition to a nine-month average immune duration, we repeated our analyses with mean immune durations between six and twelve months. As mean duration of vaccine-induced immunity decreases toward six months, the vaccination allocations minimizing mortality increasingly overrepresented elderly adults, particularly with lowest vaccine supply (10-20%). Increasing mean duration toward 12 months, more allocations overrepresenting school-age children (10-19) and working adults (20-39) minimized mortality (Figure S6). Particularly in the 10% supply, the allocations leading to minimum mortality largely do not include the 10-19 age group when the duration is seven months or less, but then primarily include this age group when the duration is ten months or higher. Under 8 to 11-month average duration, the optimal allocation under a 10% supply allocates nearly all vaccines to the 10-19 age group, but under a 12-month duration, allocating vaccines to other groups as well is optimal (though the 10-19 group still receives a majority of vaccines). The magnitude of change is shown in Figure S7, where the share that each age group comprises among the ten allocations with lowest mortality changes under assumed vaccine immunity durations. Across vaccine supplies, increasing the duration of immunity leads to the 60-69 and 70+ age groups comprising a lower share of vaccination in the most effective allocations, while the 0-9 and 10-19 groups comprise a larger share.

Vietnam has a younger population structure compared to countries in North America and Europe, with the 2019 median age in Vietnam being 31, while median ages of countries in North America and Europe range between 37 and 46 [19]. In order to see if our results favoring vaccinating younger age groups over older age groups would change if elderly adults comprised a larger share of the population, we examined different population structures. Using the United States and Vietnam as benchmarks, we repeated our analyses with populations having age structures of Vietnam, the United States, and three intermediate populations consisting of mixture distributions with weights of .25, .50, and

.75 for each country. Estimated mortality and mortality-minimizing vaccine allocations change little under different population distributions (Figures S8, S9). Under lower vaccine supplies, among the ten allocations that minimize mortality, the average share of doses allocated to different age groups remains constant (Figure S9). Under higher vaccine supplies, more changes are seen: in a population with age demographics resembling the US, the set of ten mortality-minimizing vaccine allocations allocate a higher share to the oldest age groups compared to a population with age demographics resembling Vietnam. This is likely reflective of the change in age demographics, as previously, the best vaccine allocations under higher vaccine supplies more closely resemble the population demographics compared to under lower supplies (Figures 3, 4).

**Supplementary tables**

Table S1. Parameters assumed, taken from literature, and estimated for mathematical model, including references for values or methods of estimation.

| Parameter | Value | Reference or method of estimation |
|---|---|---|
| transmission parameter for A/H1 | Estimated in parameter search | Middle value after drawing<br>1. $\mu_\beta \sim U(0.15, 0.70)$<br>2. 3 Draws of $N(\mu_\beta, 0.01)$ |
| transmission parameter for A/H3 | Estimated in parameter search | Highest value after drawing<br>1. $\mu_\beta \sim U(0.15, 0.70)$<br>2. 3 Draws of $N(\mu_\beta, 0.01)$ |
| transmission parameter for B | Estimated in parameter search | Lowest value after drawing<br>1. $\mu_\beta \sim U(0.15, 0.70)$<br>2. 3 Draws of $N(\mu_\beta, 0.01)$ |
| Times of peak transmission | Estimated in parameter search | Drawn from distributions estimated previously [2]; spacings between peaks are distributed N(376, 150); (sub)type selected randomly for each peak |
| Cross immunity across (sub)types | A/H1 and A/H3: 30%<br>A/H1 and B: 27%<br>A/H3 and B: 17%<br>Assumed symmetric | [1] |
| Duration of increased transmission (days) | Estimated in parameter search | Drawn from U(45, 180) |
| Magnitude of increased transmission (multiplier) | Estimated in parameter search | Drawn from U(0, 0.5) |
| Duration between case importations | Estimated in parameter search | Drawn from U(90, 365) |
| Hospitalization fraction among cases, all (sub)types | 0-9: 0.0048<br>10-19: 0.0033<br>20-29: 0.0056<br>30-39: 0.0056<br>40-49: 0.0056<br>50-59: 0.0106<br>60-69: 0.0460<br>70+: 0.0909 | [4] |
| Death fraction among hospitalizations, all (sub)types | 0-9: 0.0094<br>10-19: 0.0125<br>20-29: 0.0289 | [4] |

| | | |
|---|---|---|
| | 30-39: 0.0289<br>40-49: 0.0289<br>50-59: 0.0574<br>60-69: 0.0780<br>70+: 0.1041 | |
| Vaccine effectiveness | 0.5 | Assumed 0.5 for effectiveness against infection, hospitalization if infected, and death if hospitalized [13–17] |
| Duration of infection-induced immunity (days) | Estimated in parameter search | Drawn from U(180, 1460) |
| Duration of vaccine-induced immunity | 270 days | Assumed [18] |
