## Supplementary figures and images for "Influenza vaccination allocation in tropical settings under constrained resources"

### Figure S1

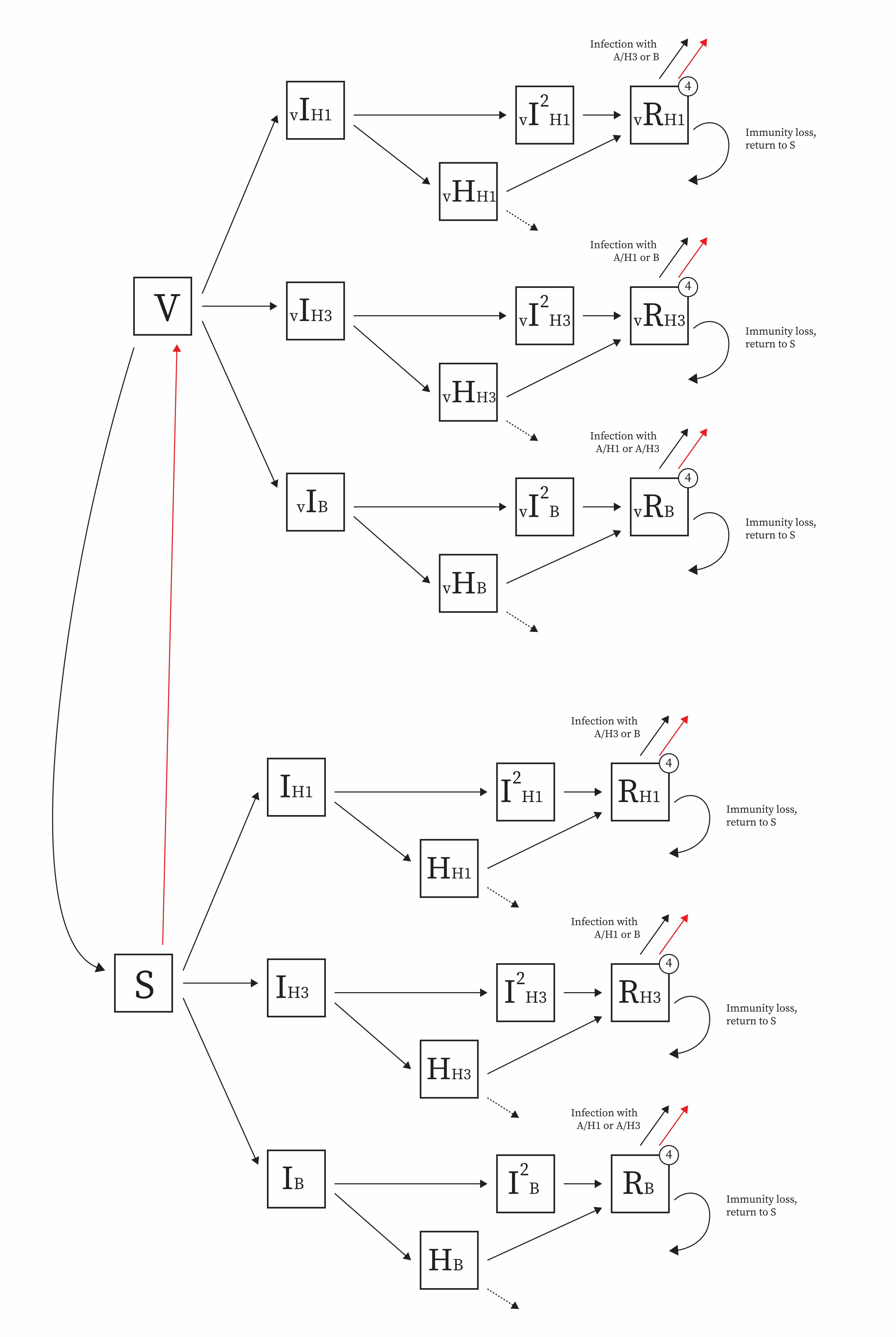

### Figure S2

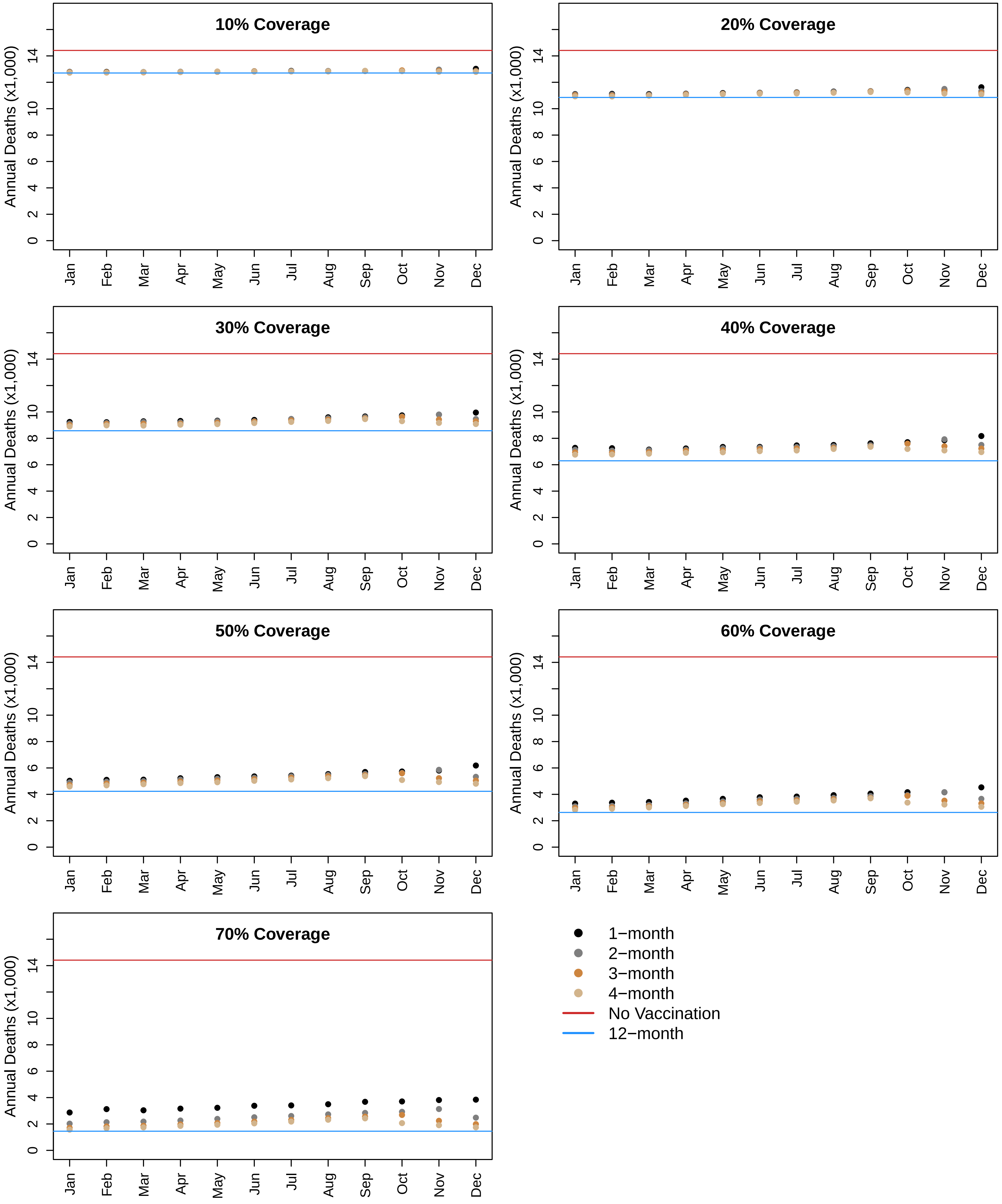

### Figure S3

#### 10% Supply

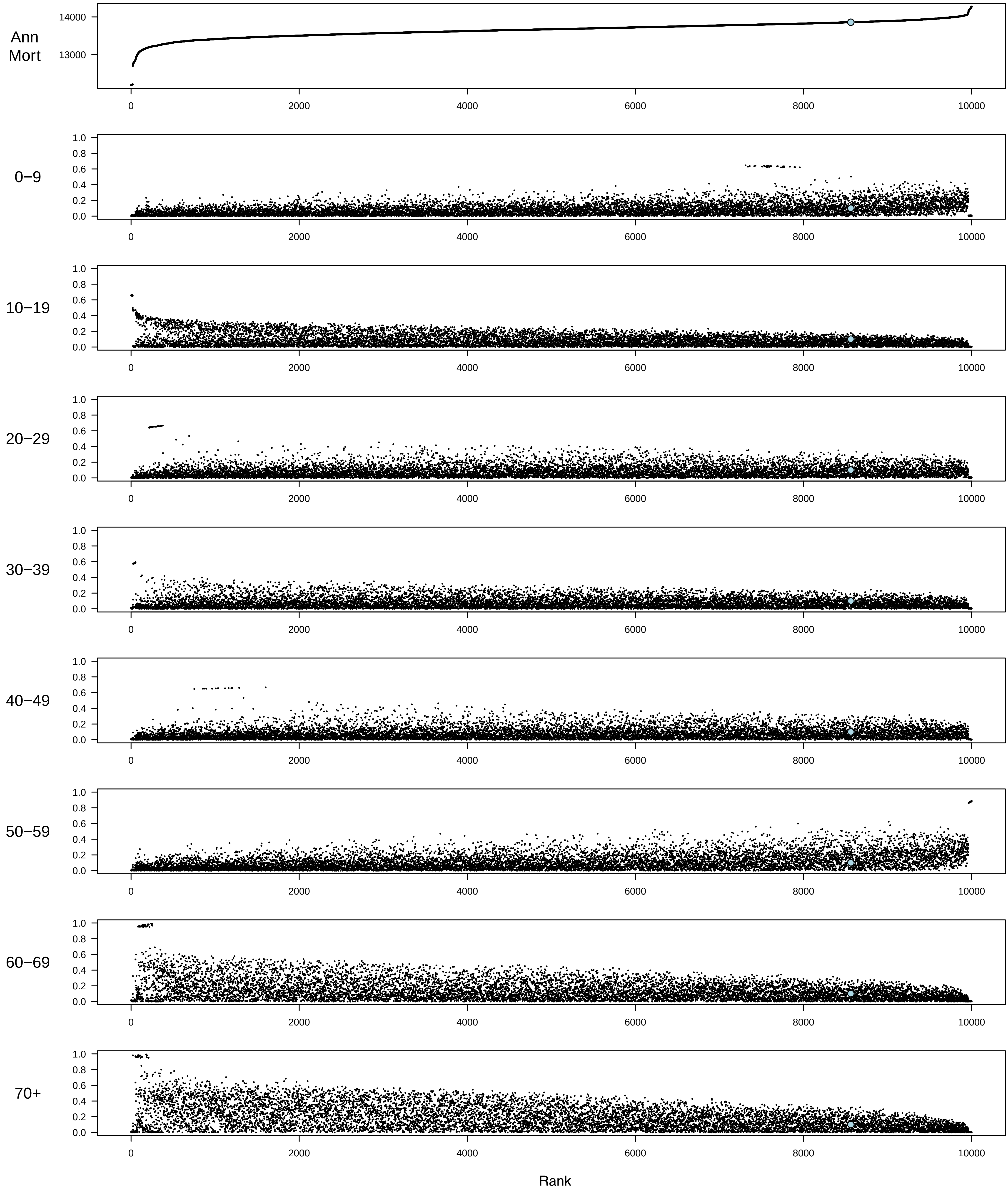

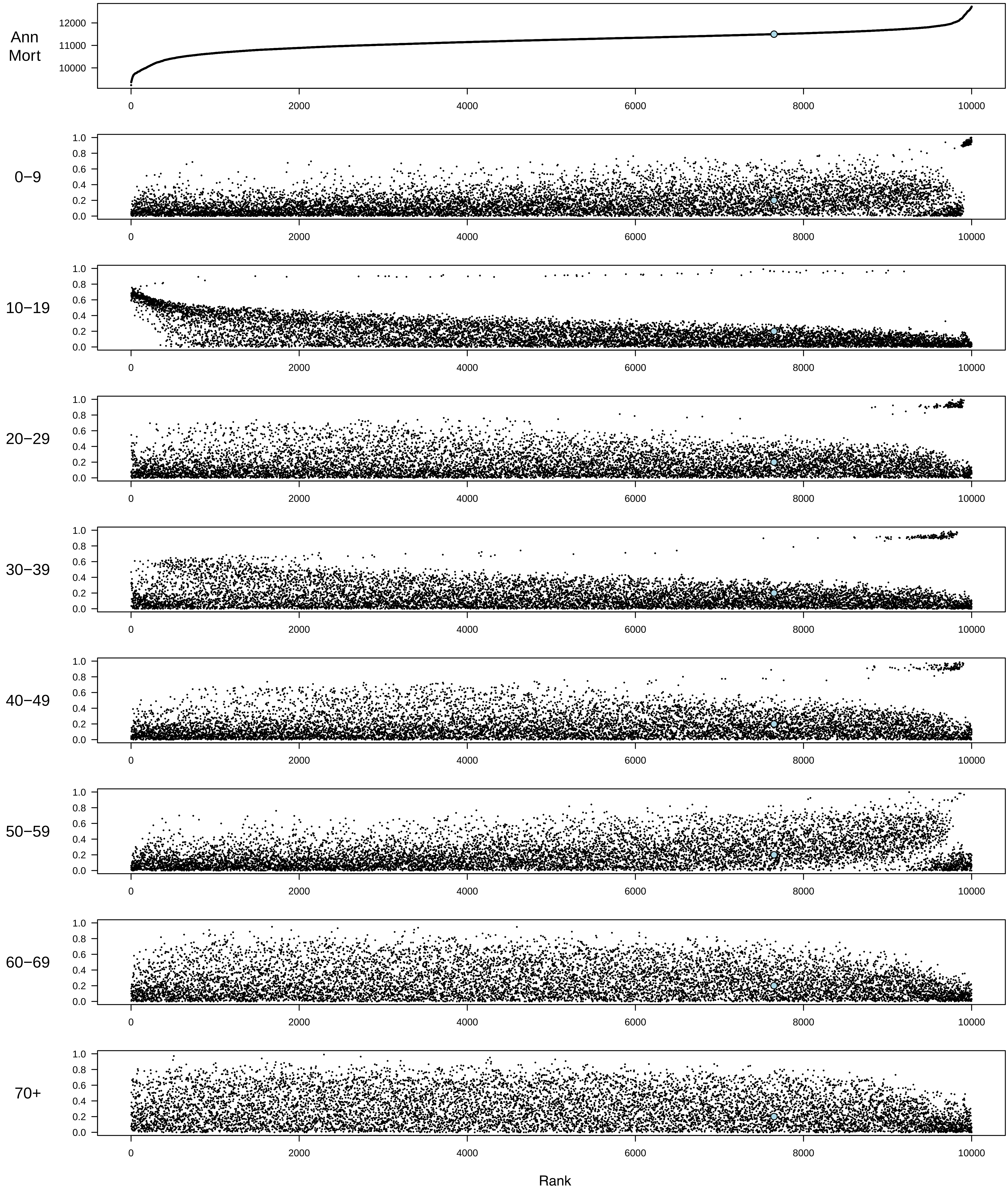

#### 30% Supply

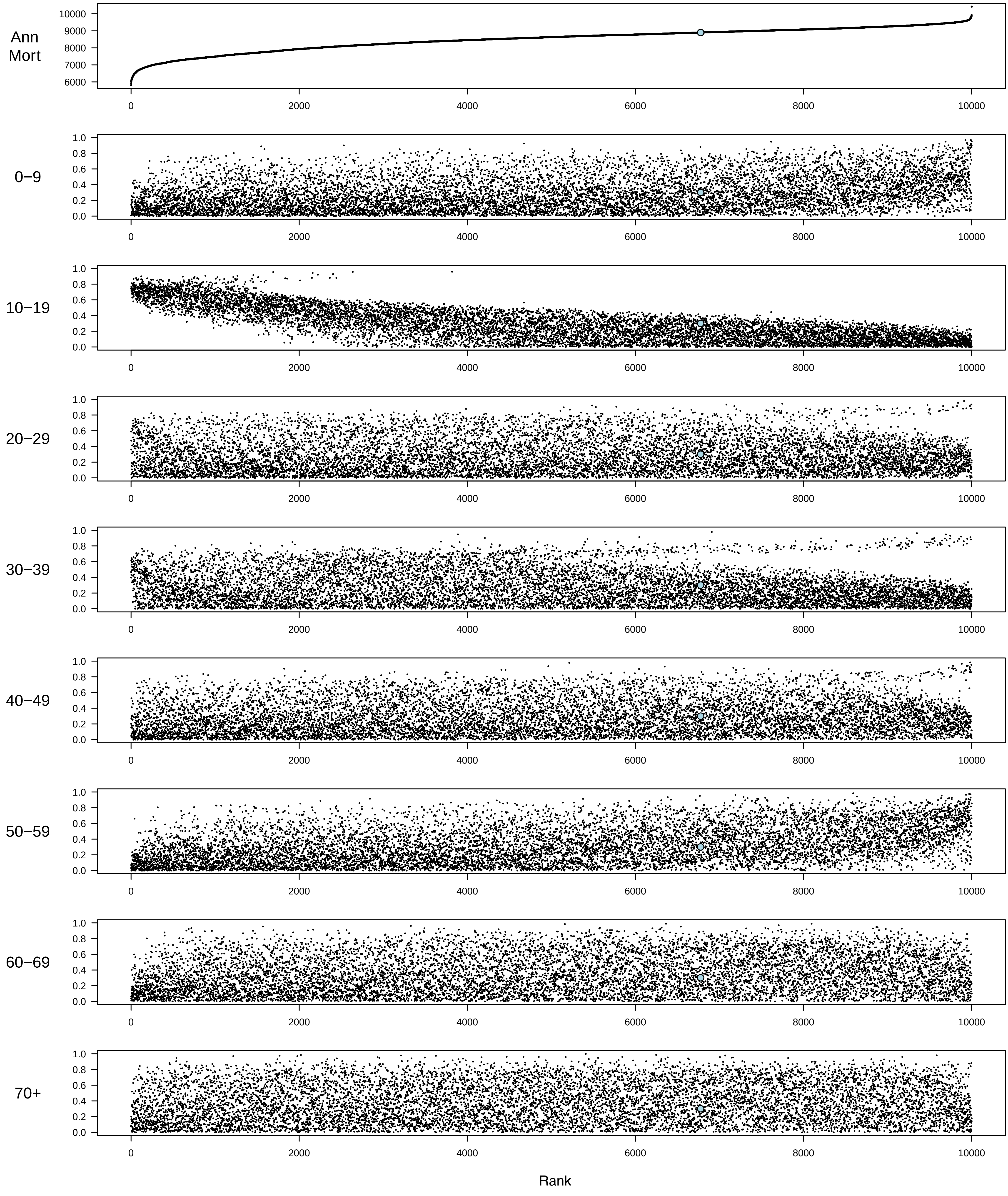

#### 40% Supply

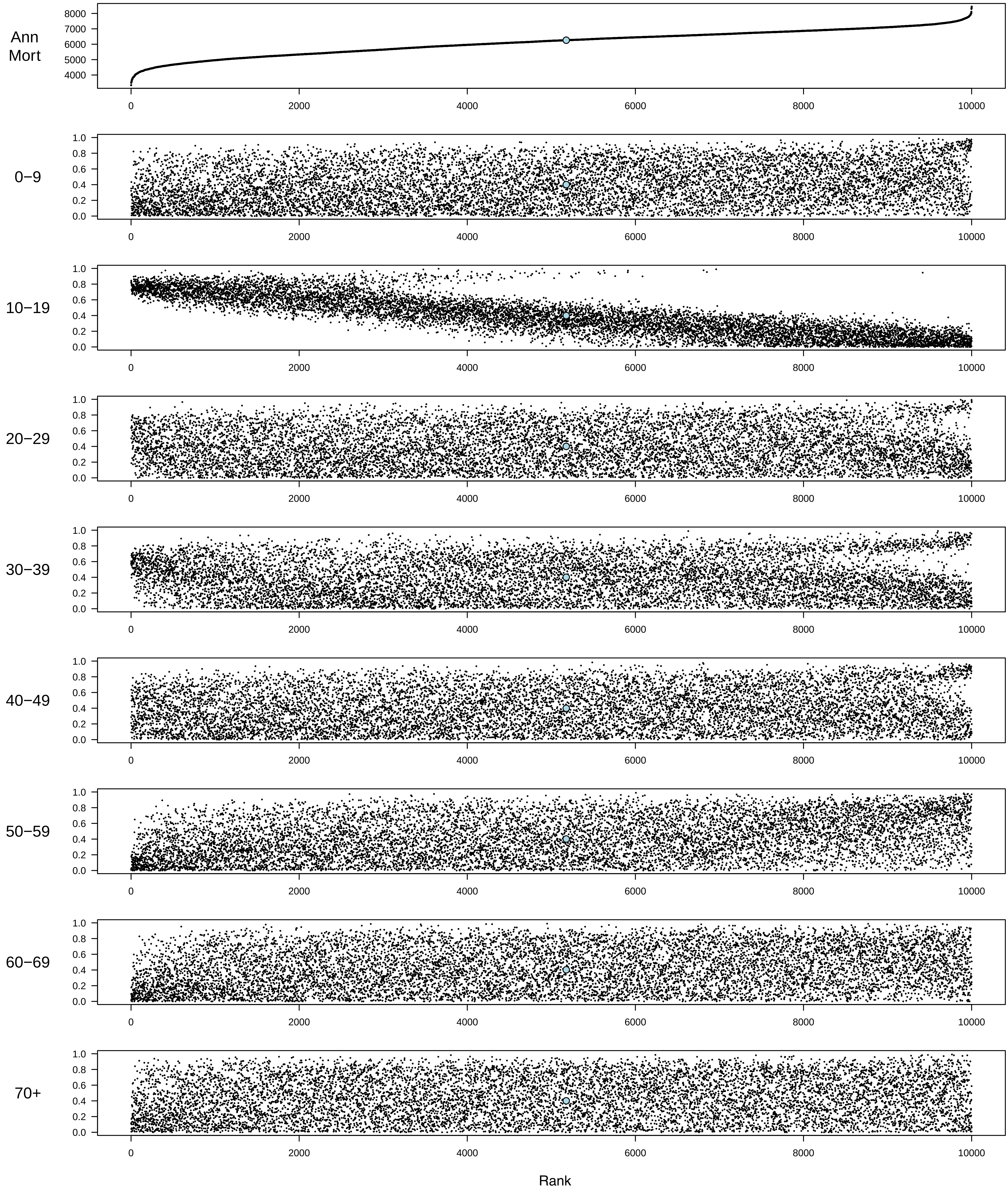

#### 50% Supply

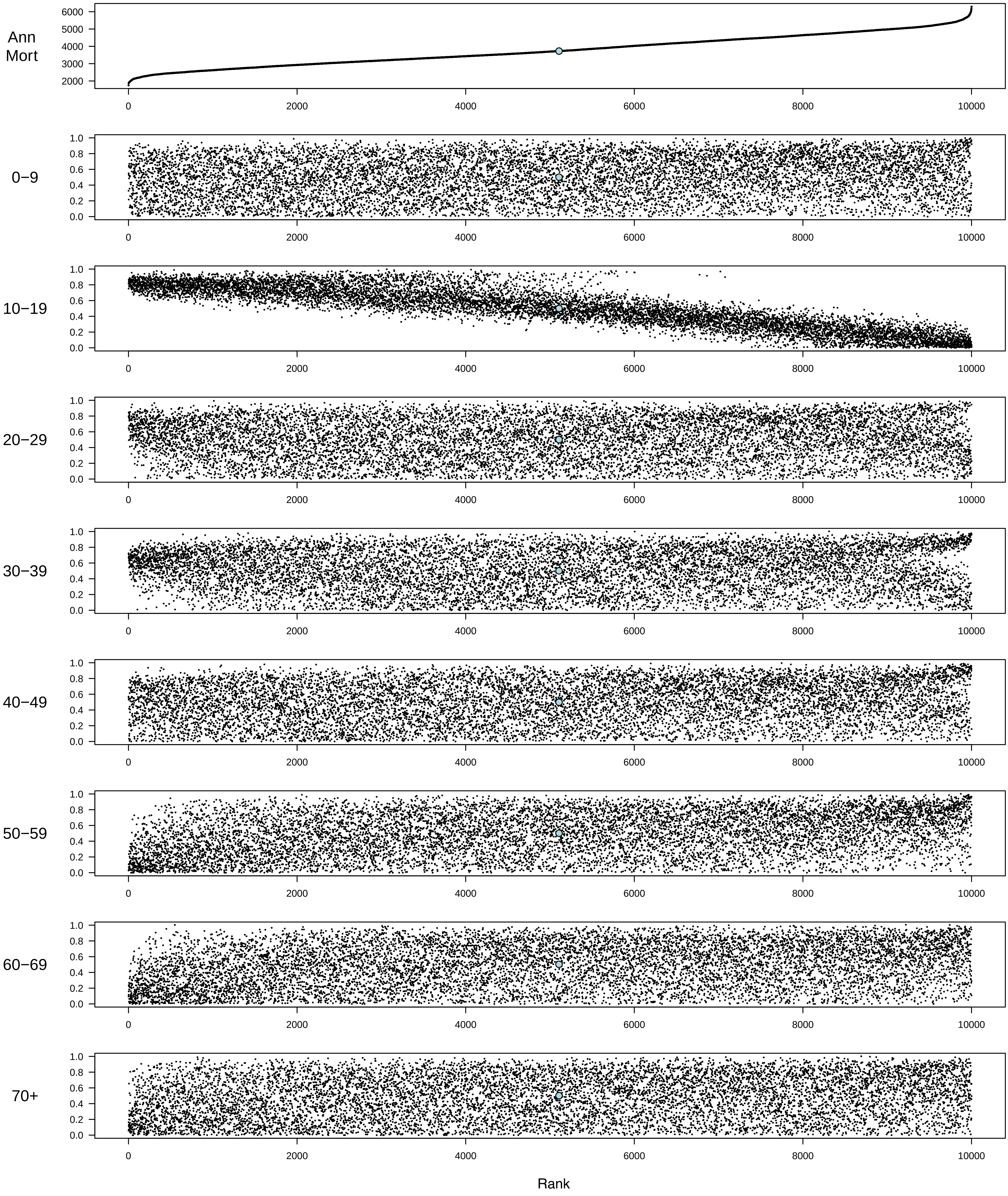

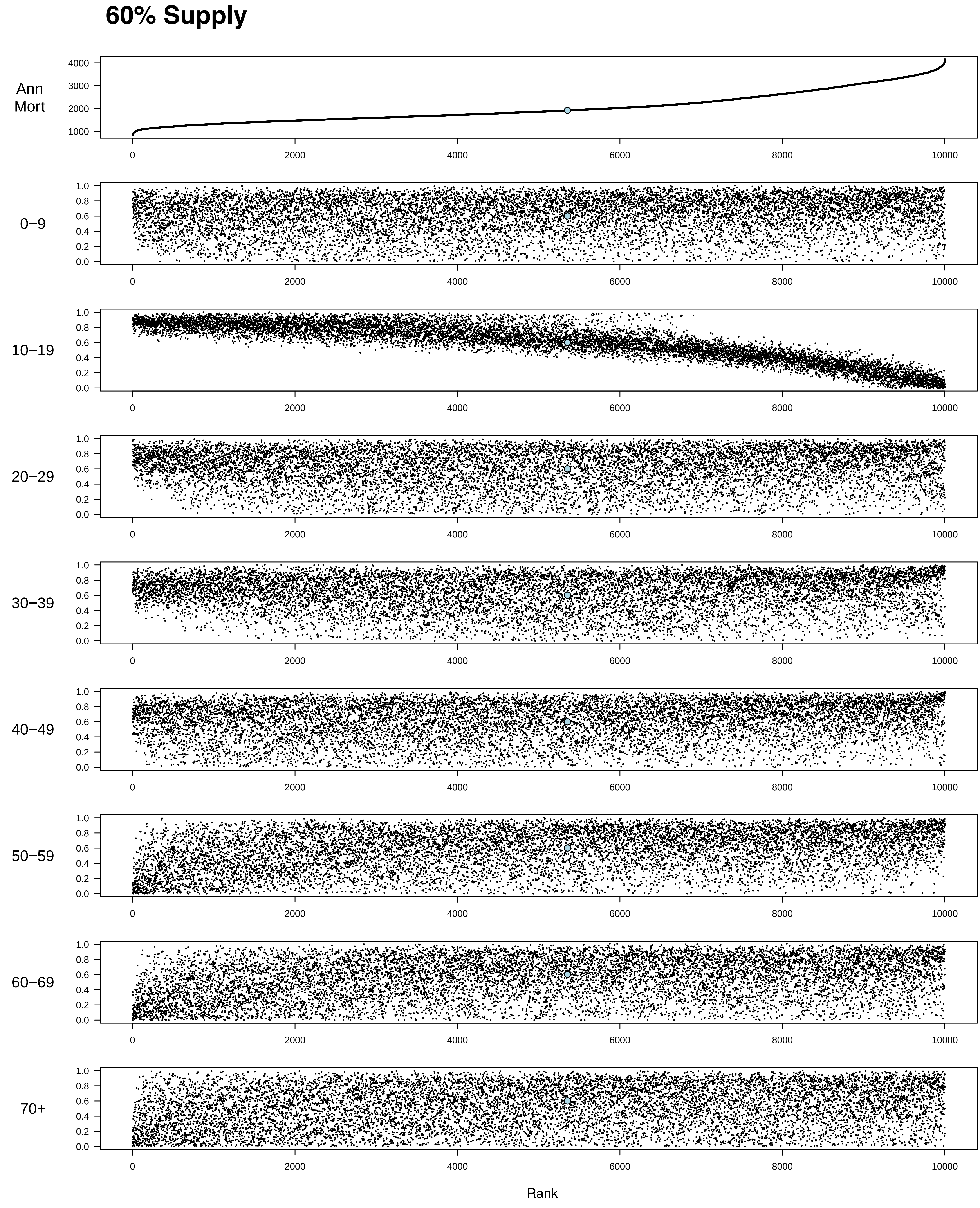

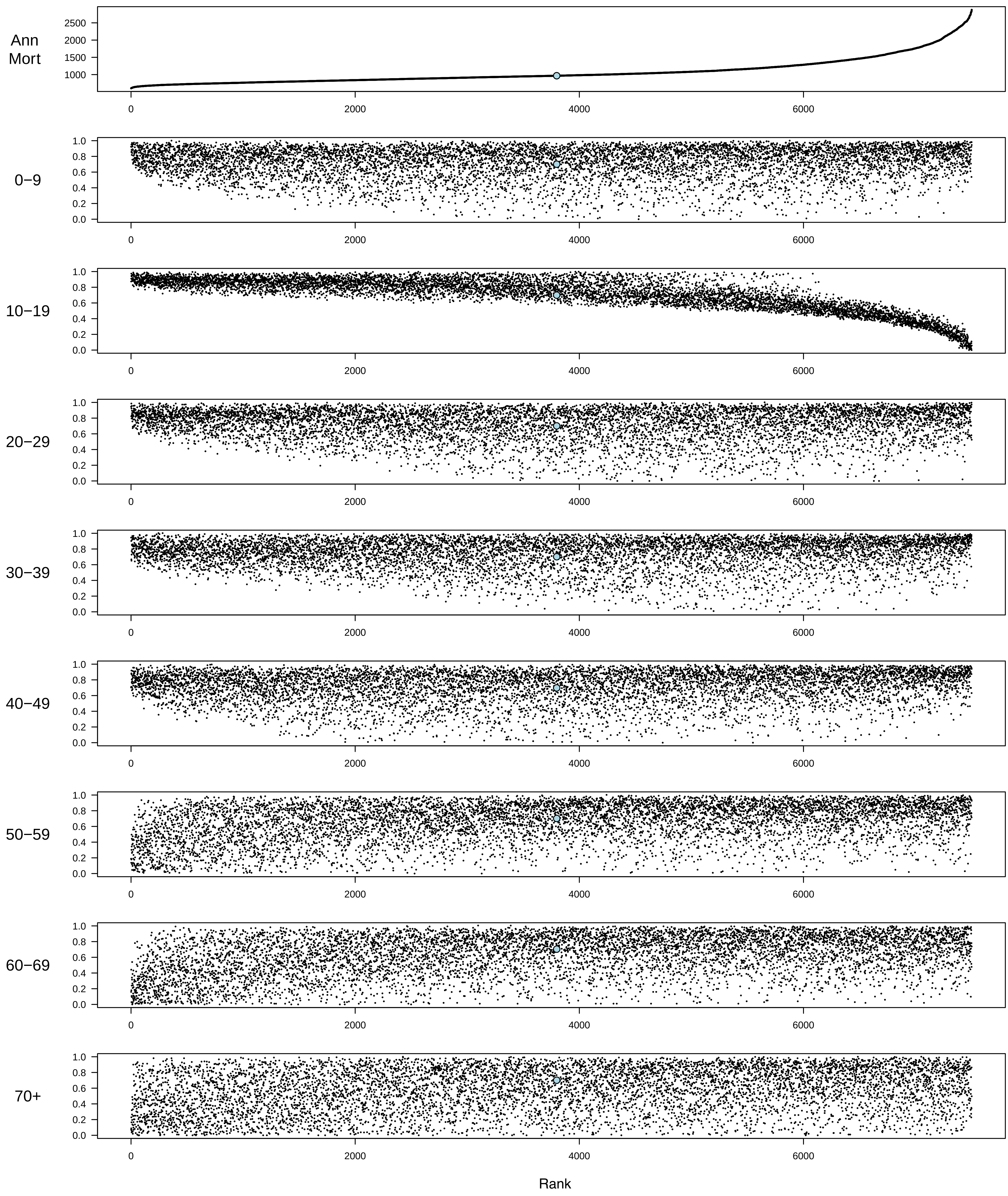

#### 80% Supply

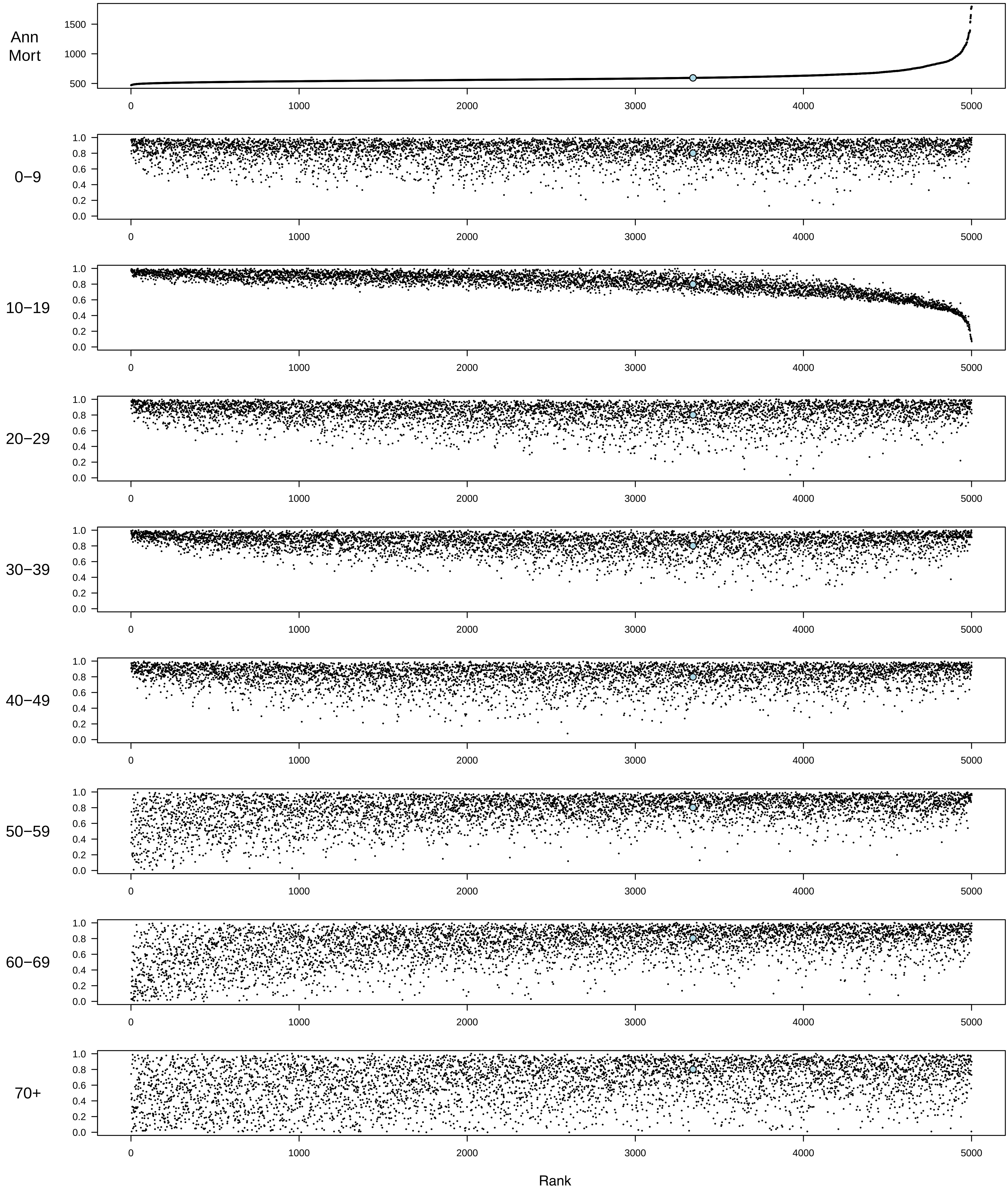

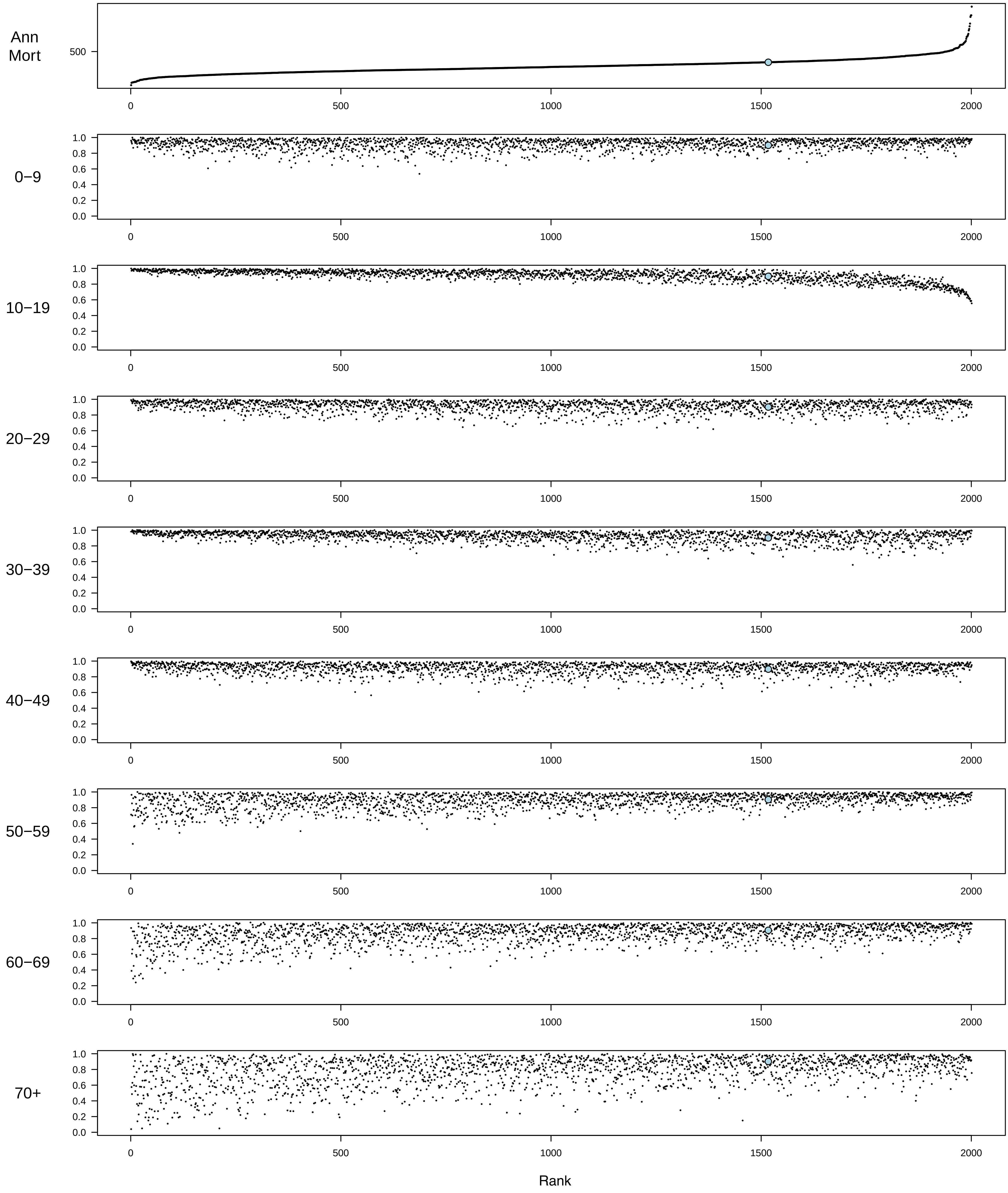

### Figure S5

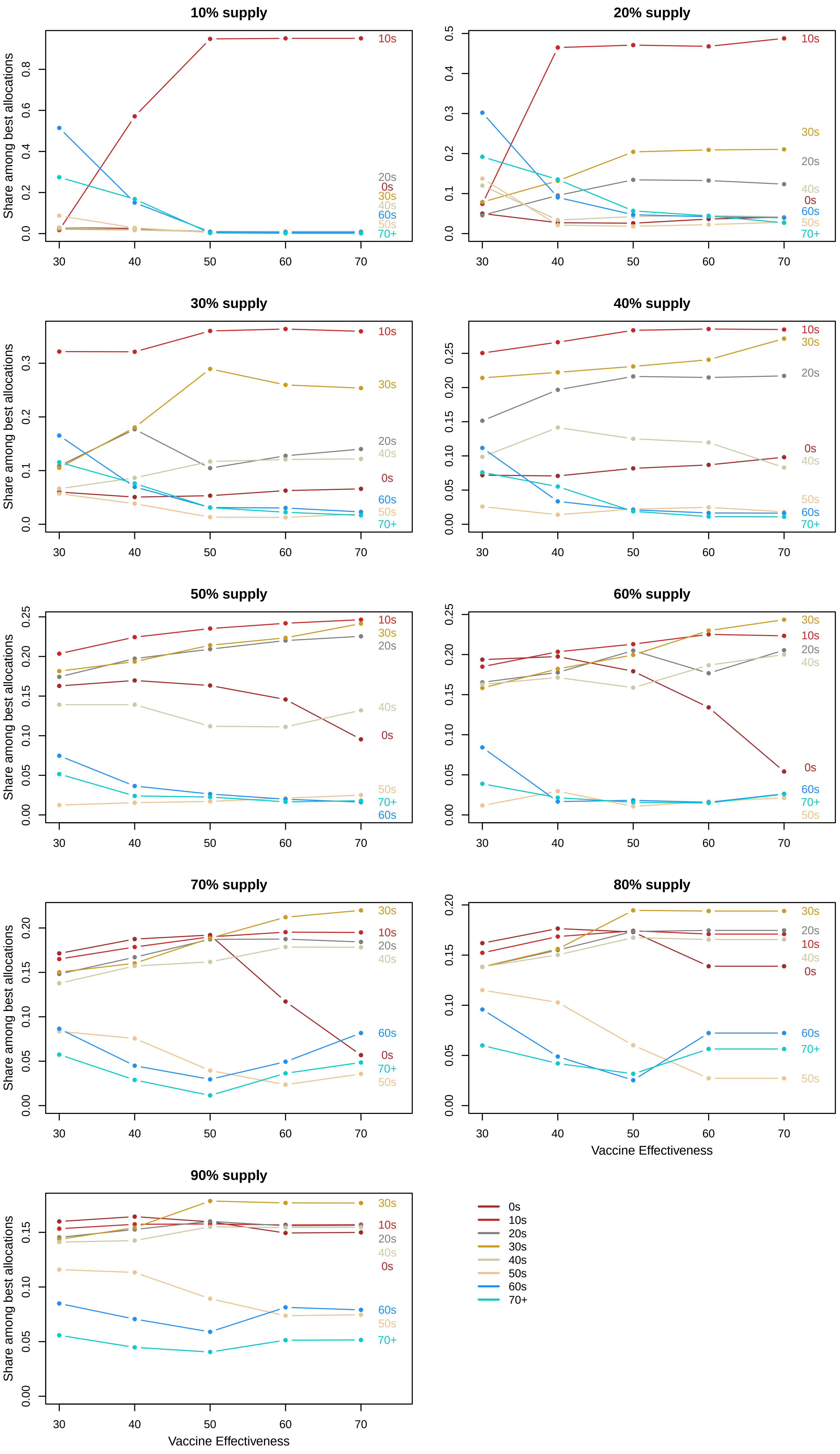

### Figure S7

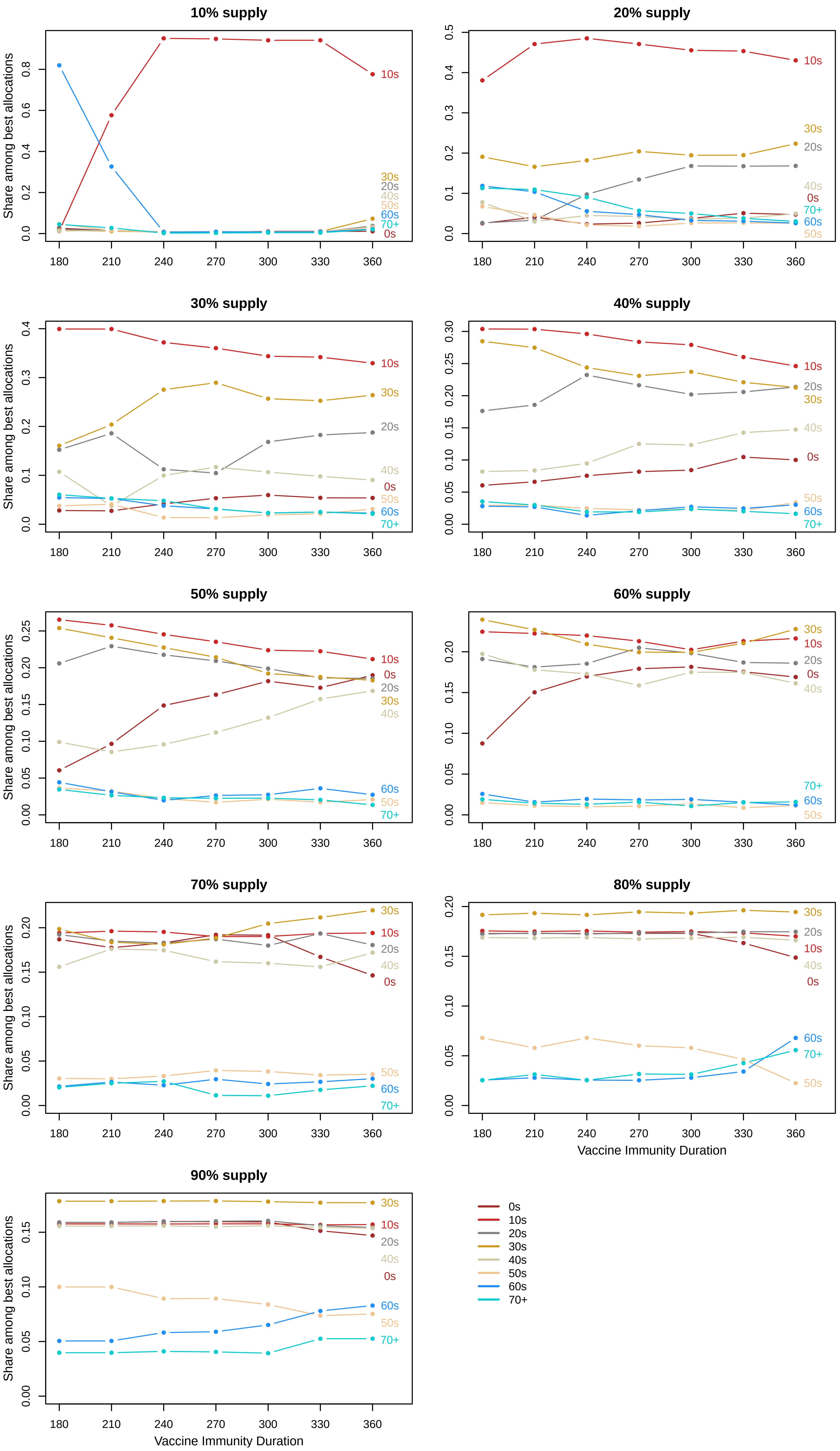

### Figure S9

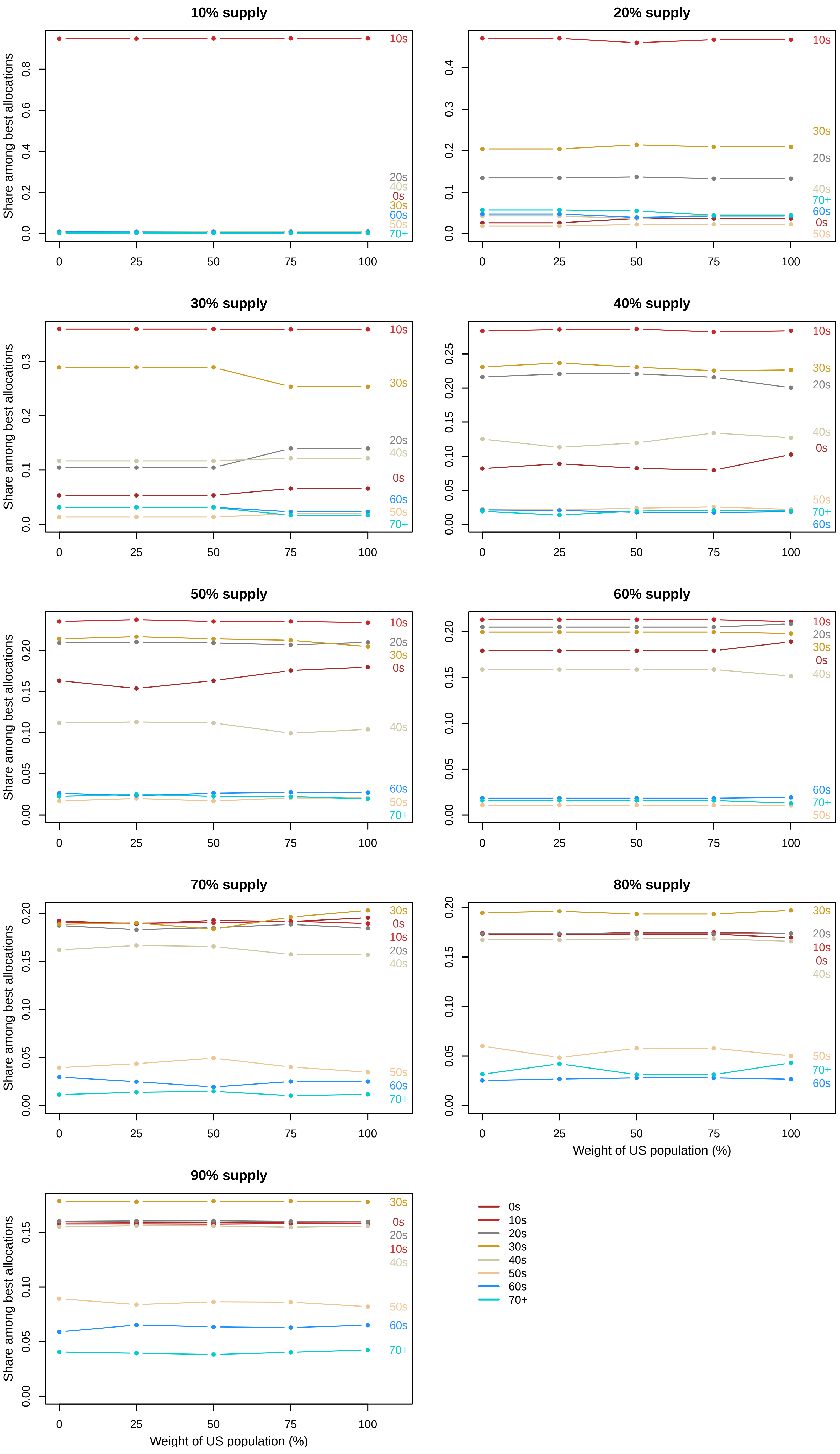
