## Supplementary material for "Influenza vaccination allocation in tropical settings under constrained resources": Figure S4

### 30% Vaccine Effectiveness

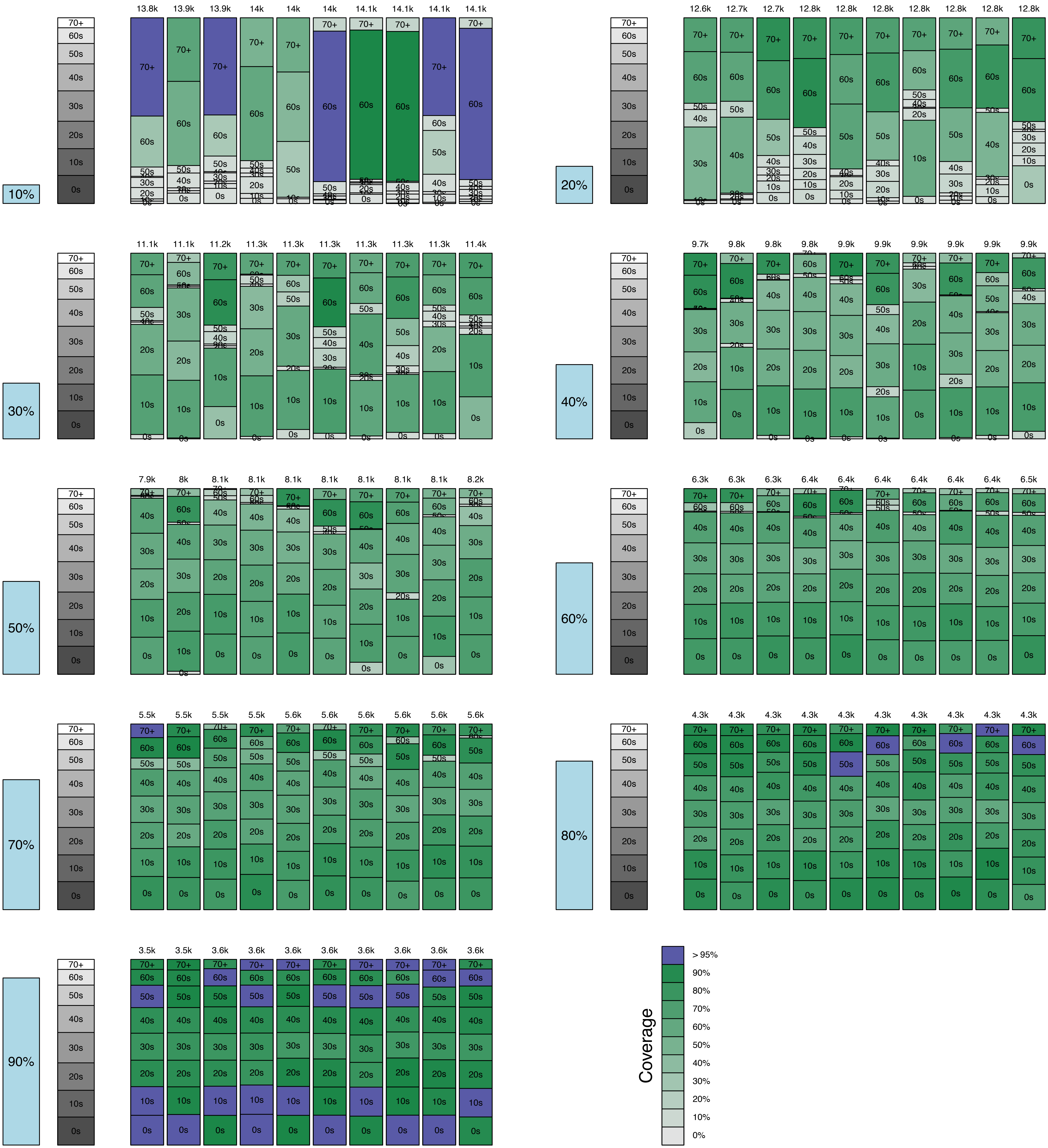

### 40% Vaccine Effectiveness

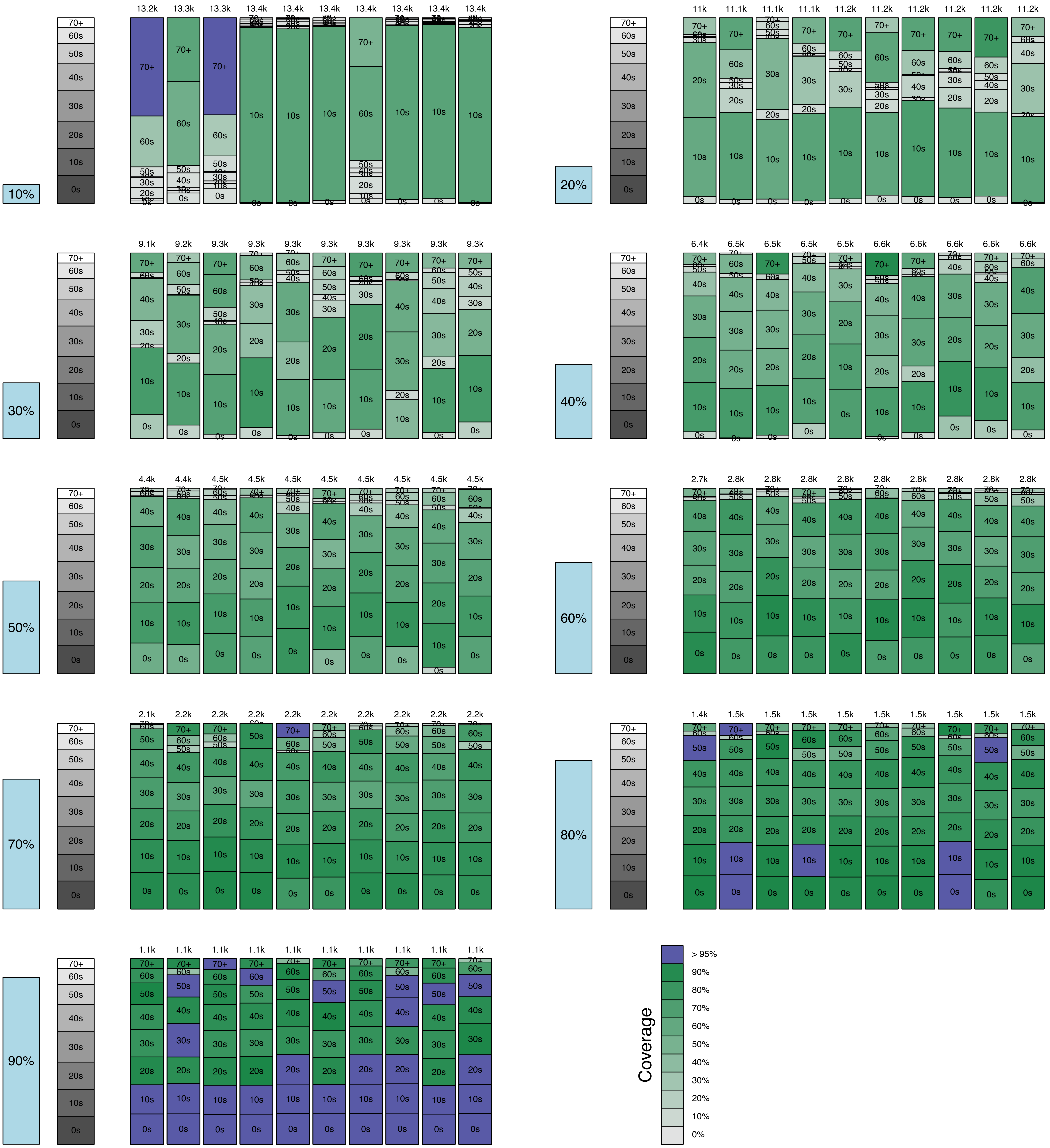

### 50% Vaccine Effectiveness

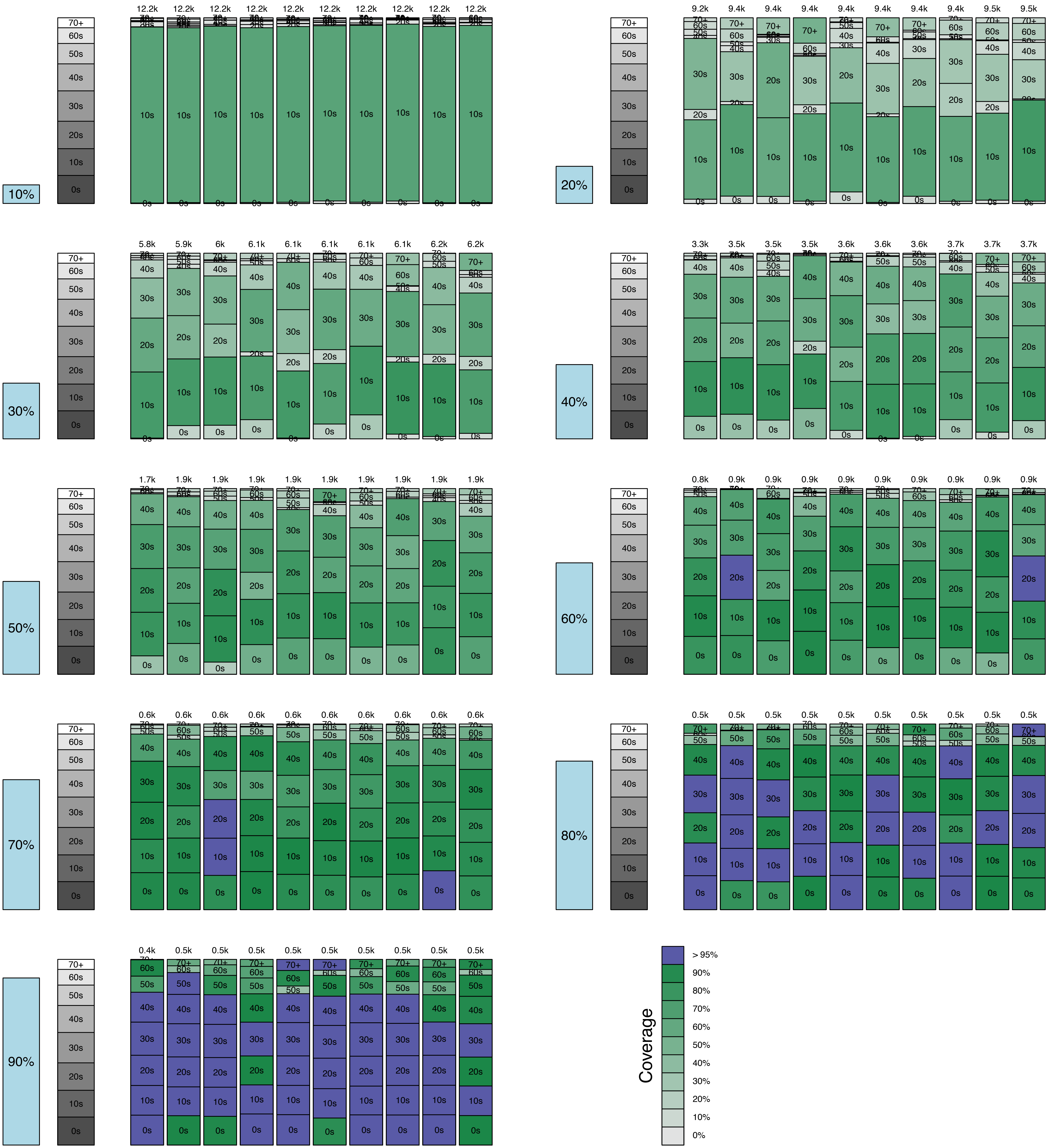

### 60% Vaccine Effectiveness

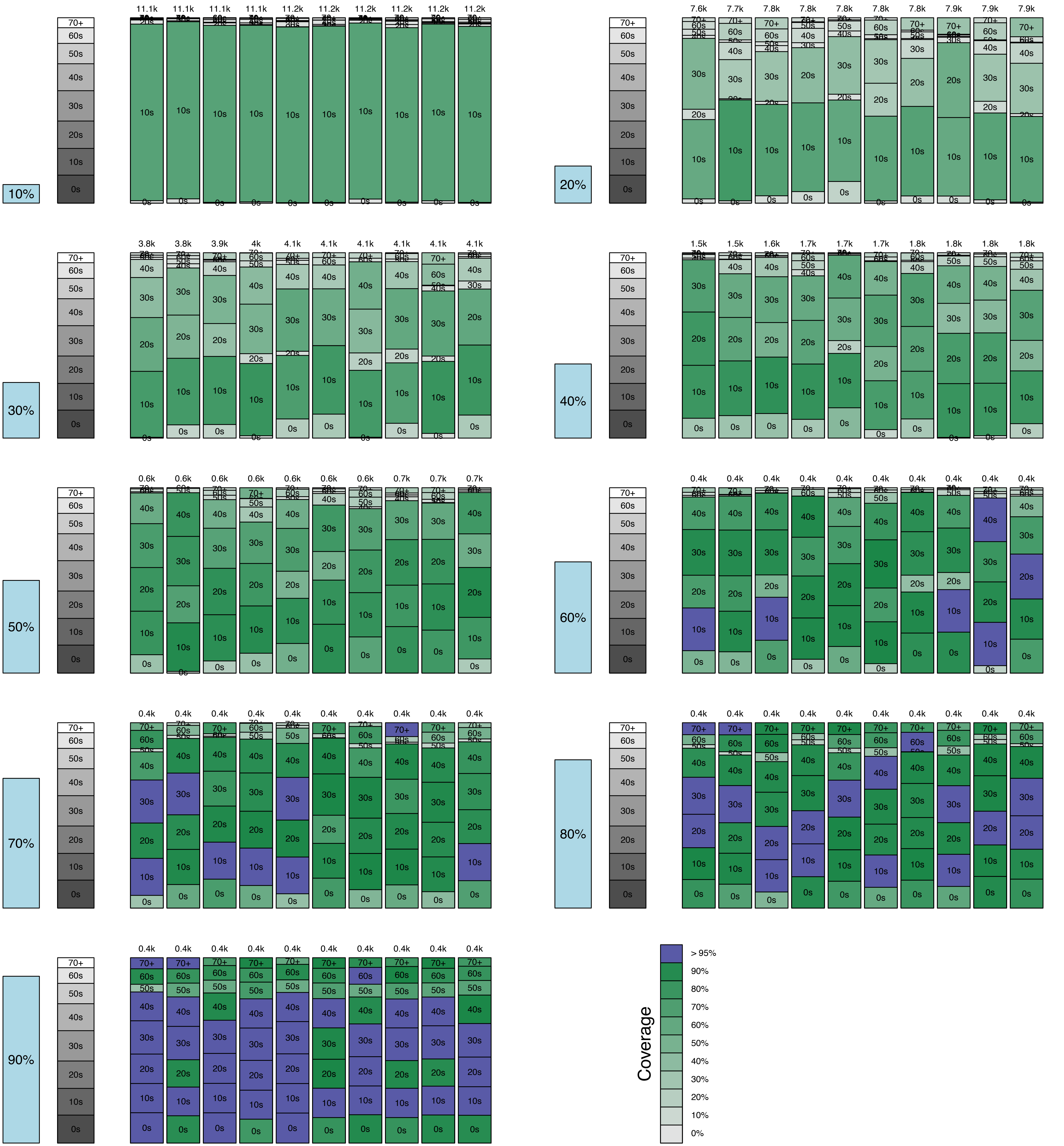

### 70% Vaccine Effectiveness

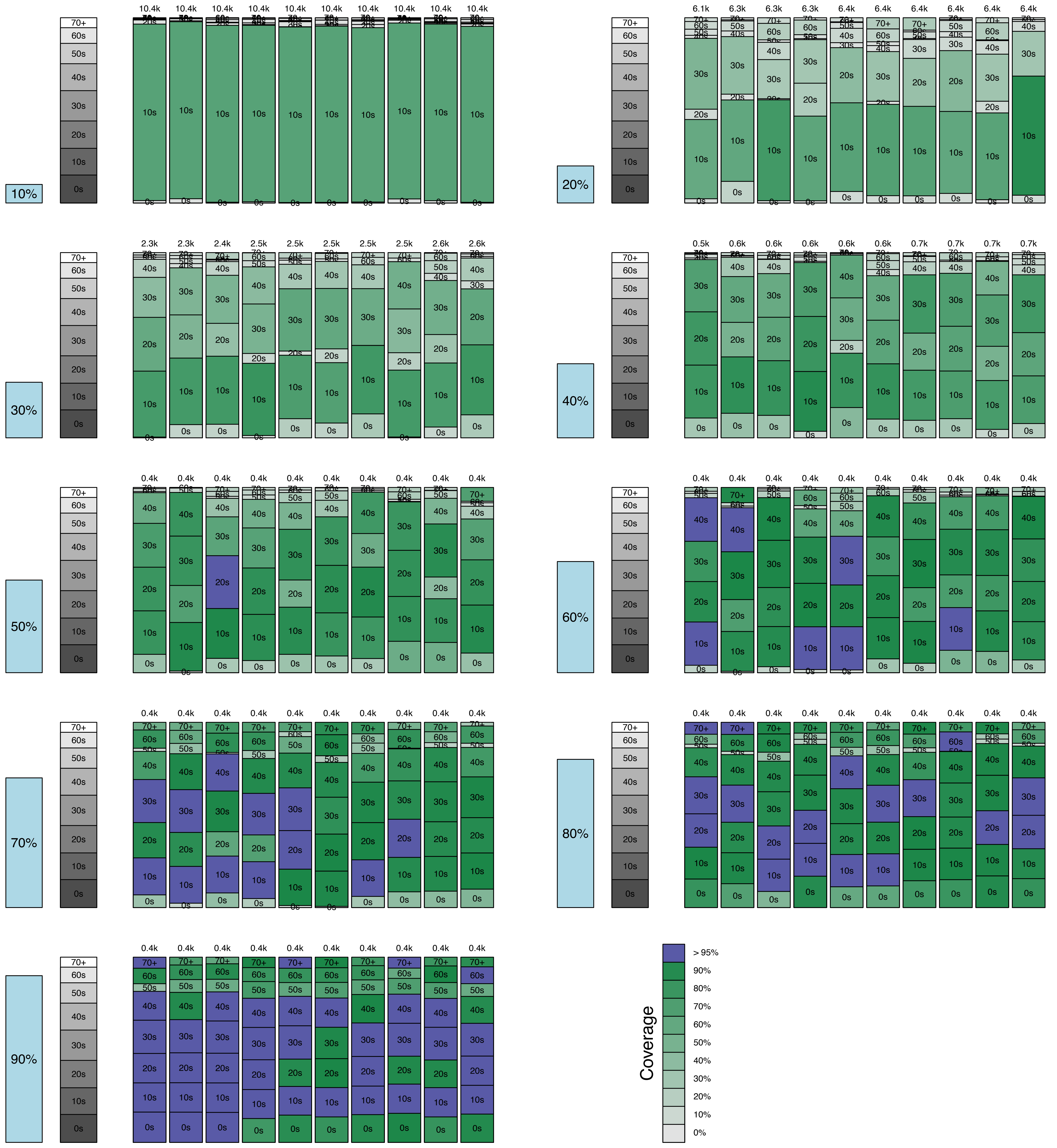
