## Supplementary material for "Influenza vaccination allocation in tropical settings under constrained resources": Figure S6

### 180-day Immune Duration

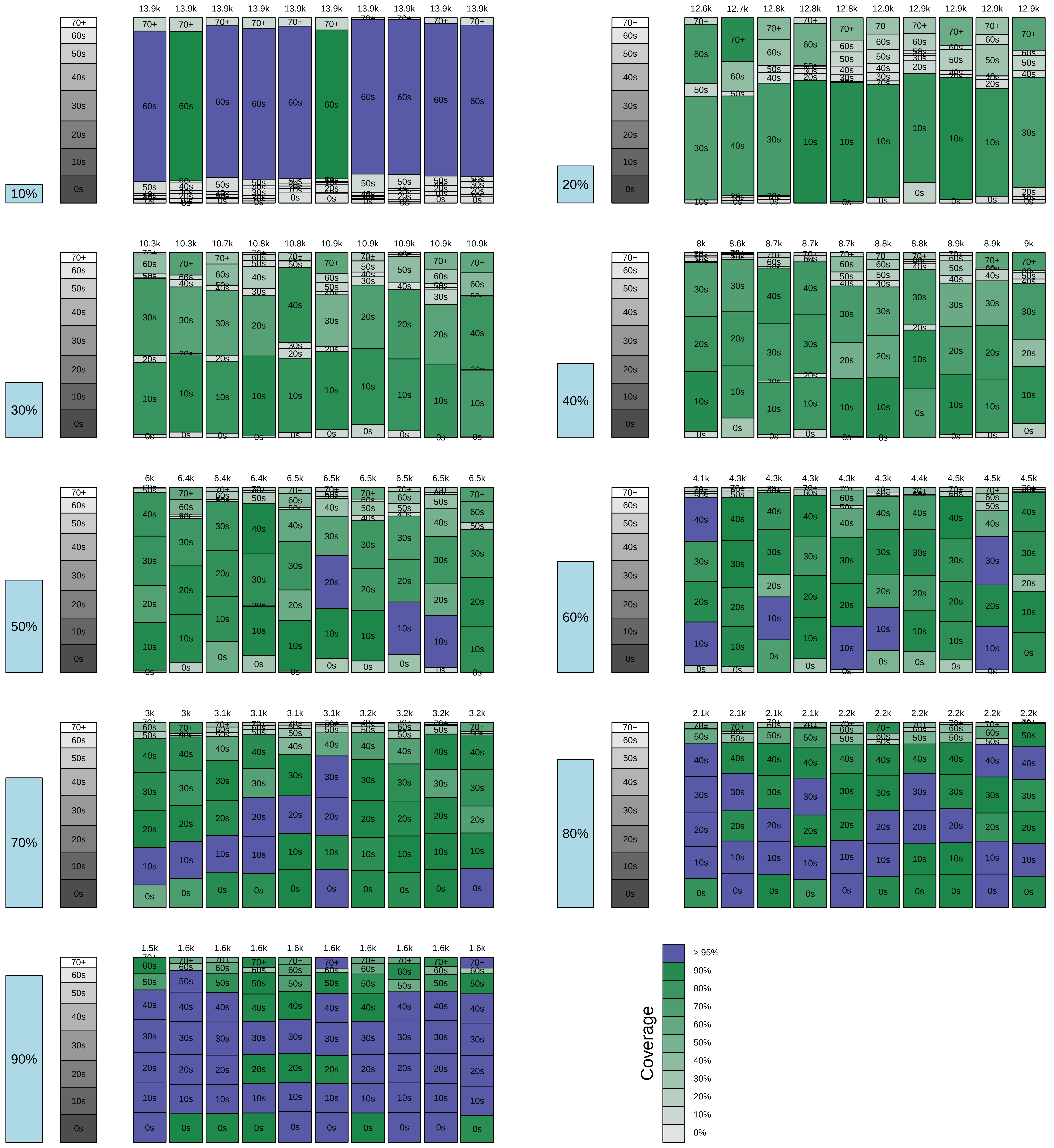

### 210-day Immune Duration

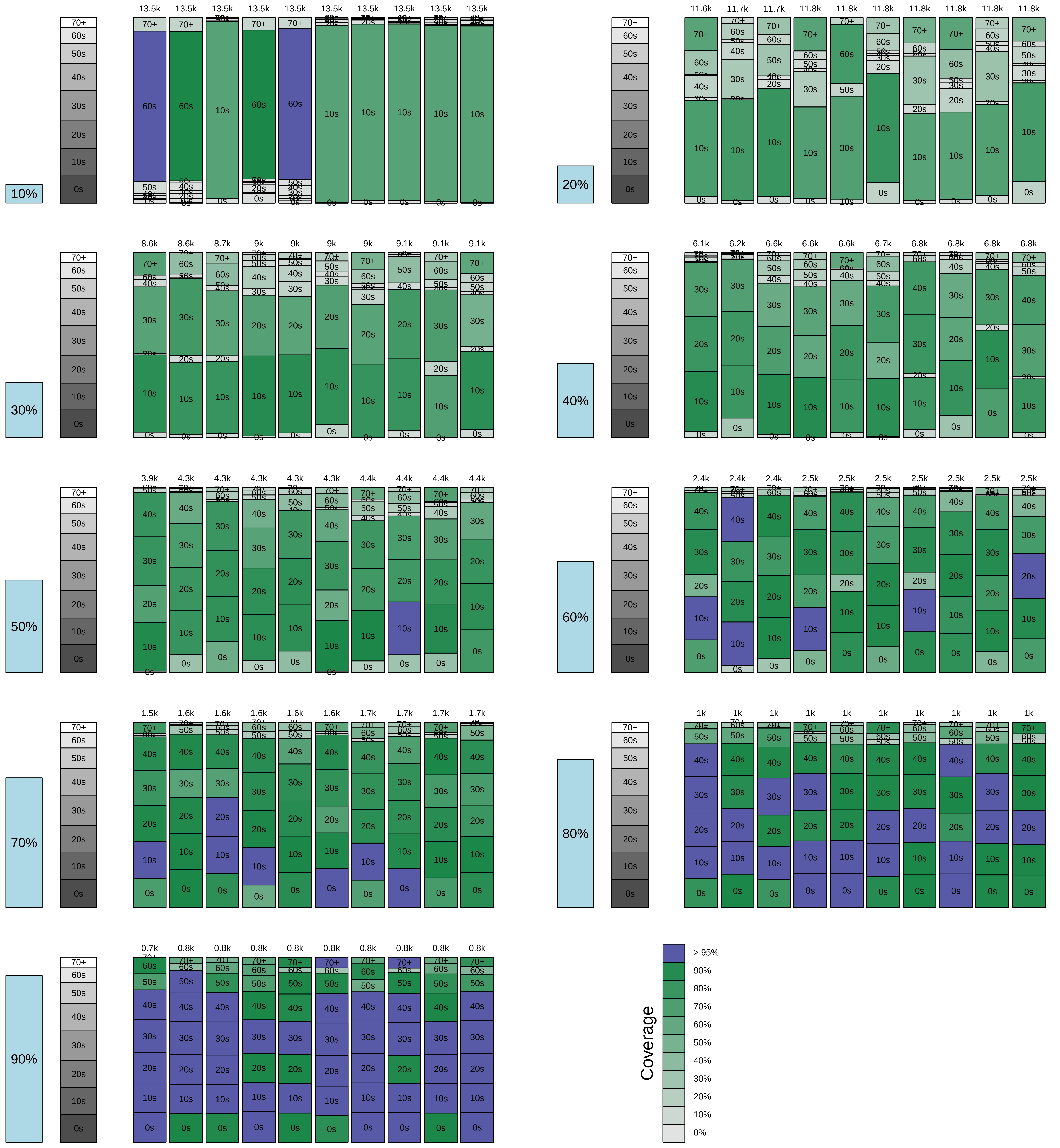

### 240-day Immune Duration

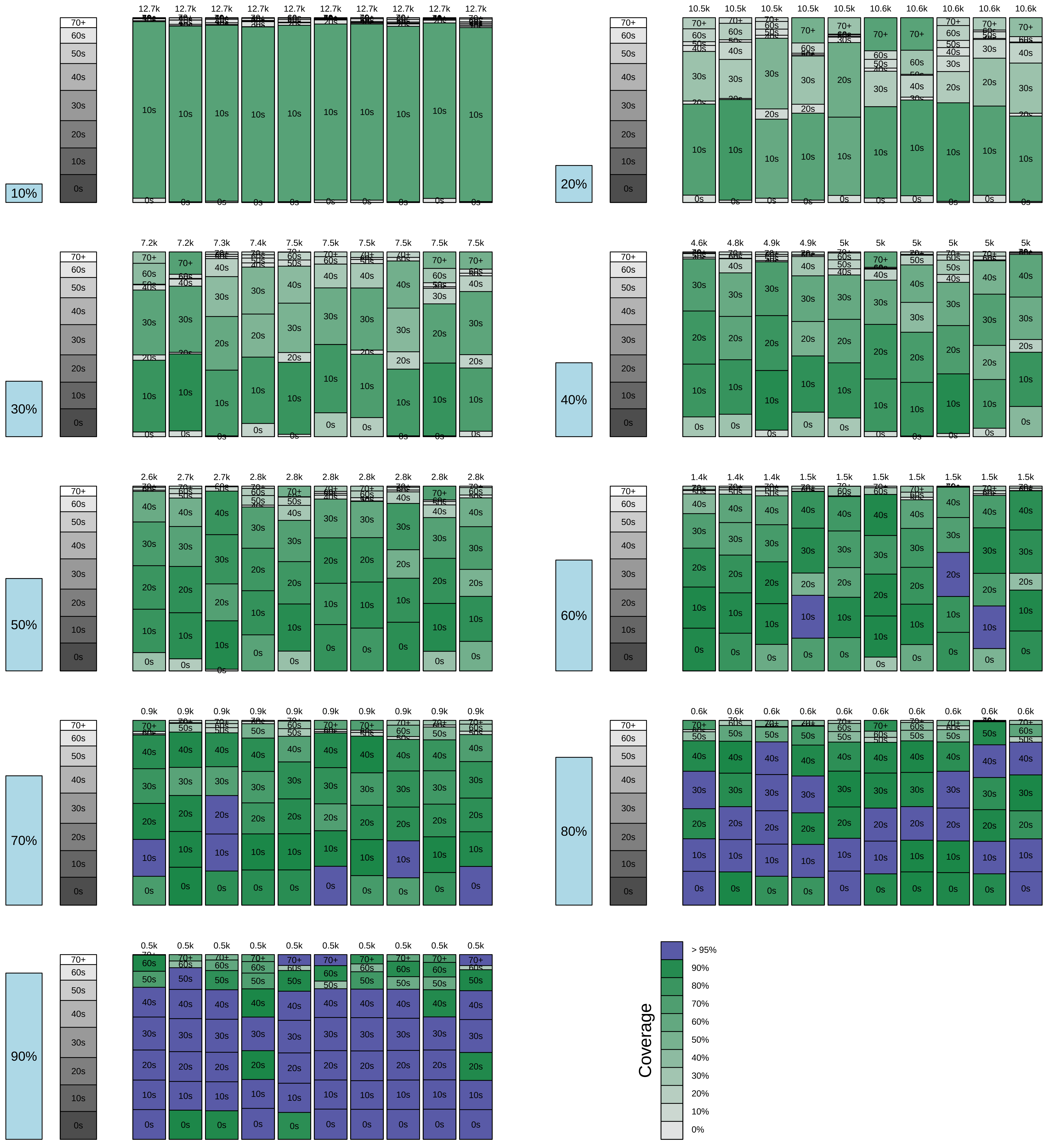

### 270-day Immune Duration

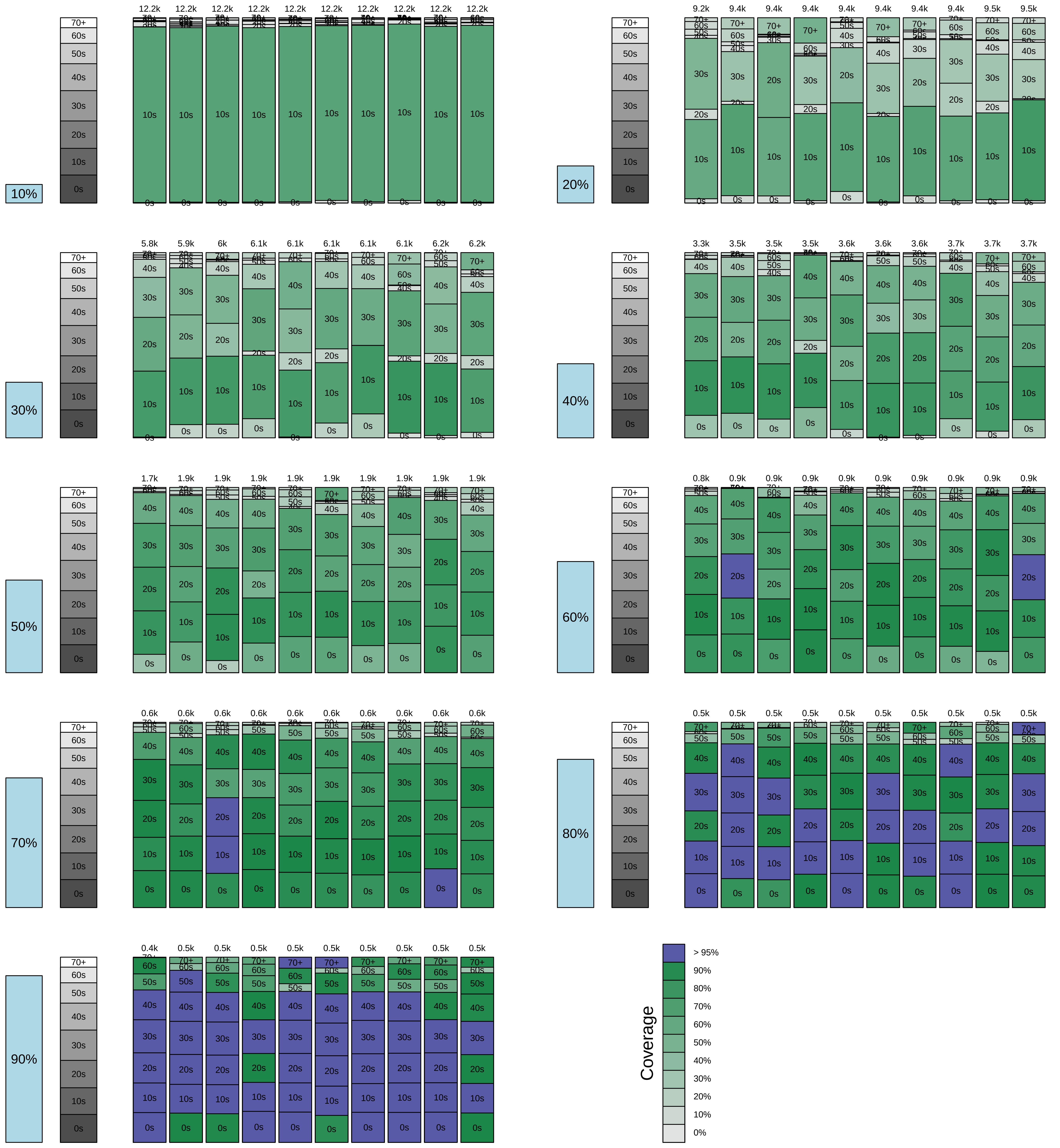

### 300-day Immune Duration

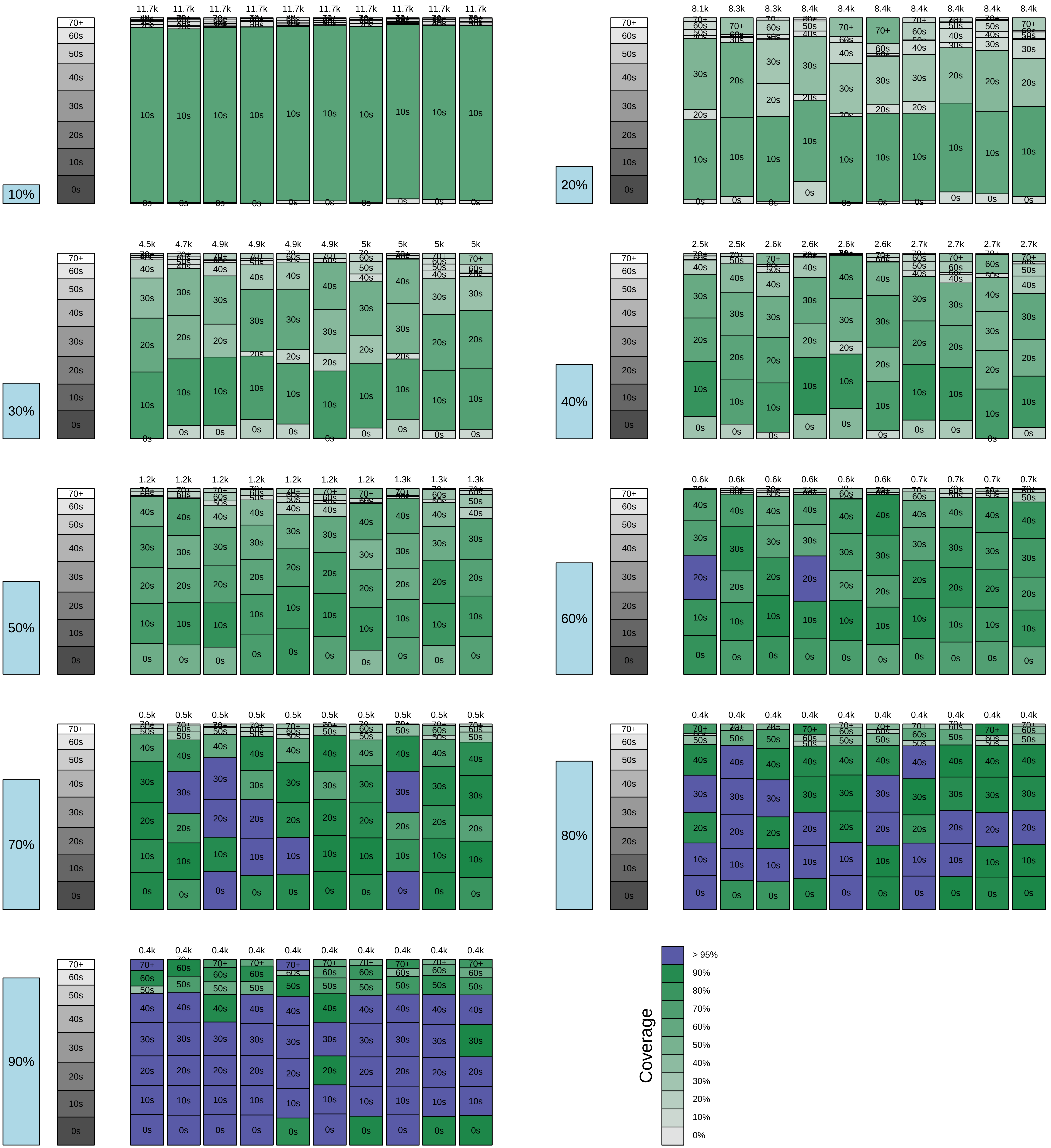

### 330-day Immune Duration

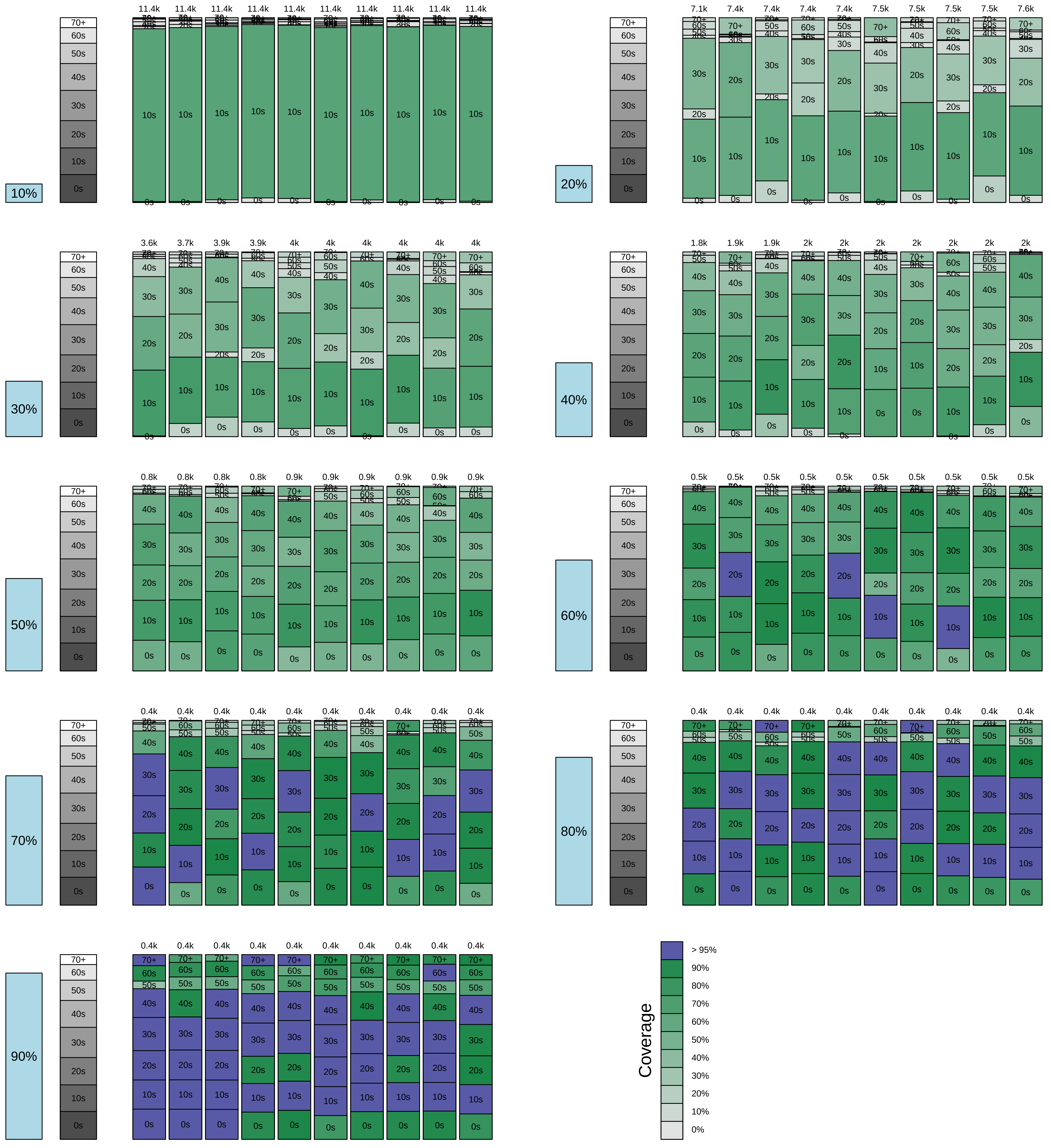

### 360-day Immune Duration

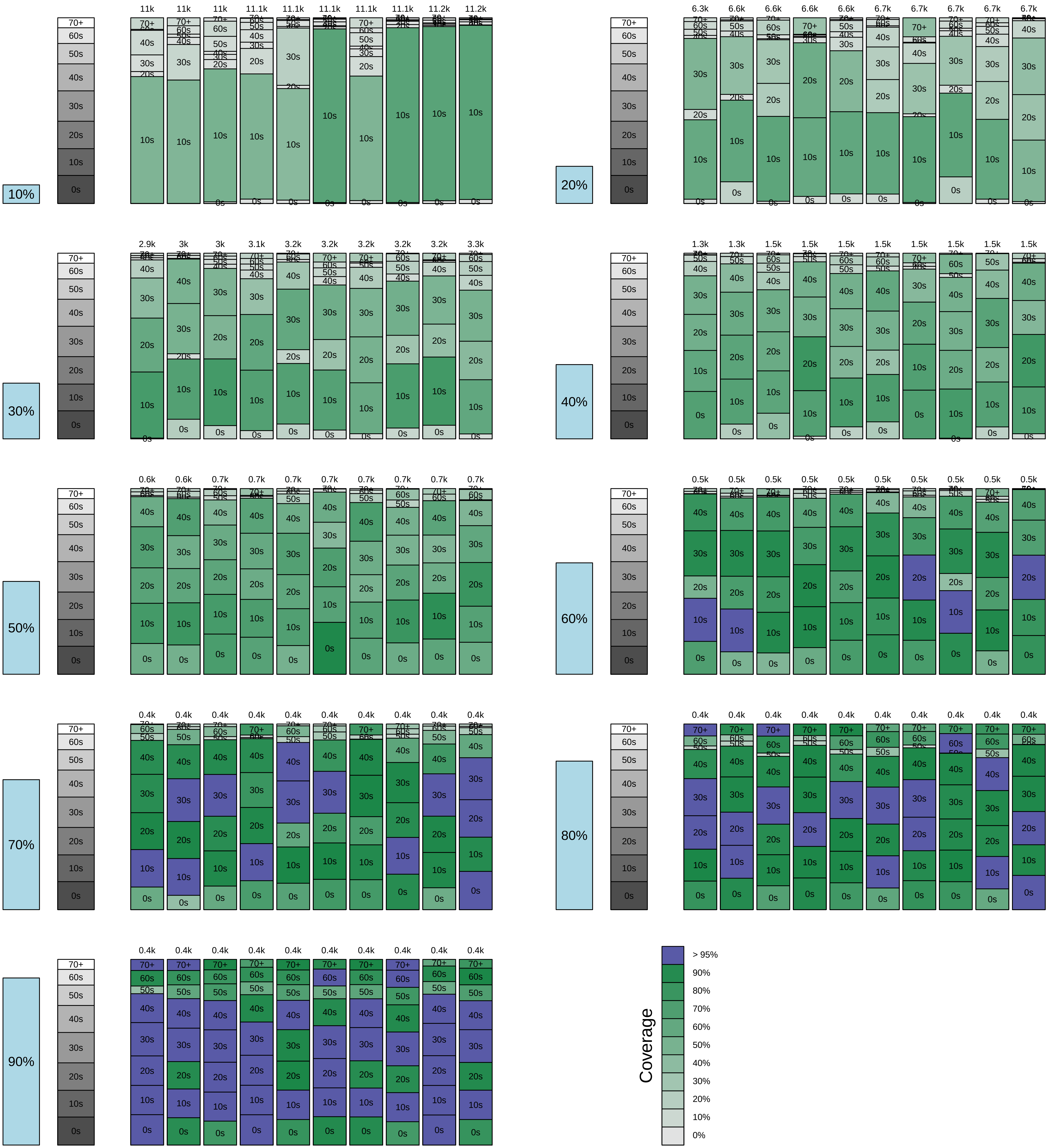
