## Supplementary material for "Influenza vaccination allocation in tropical settings under constrained resources": Figure S8

### 100% VN Population

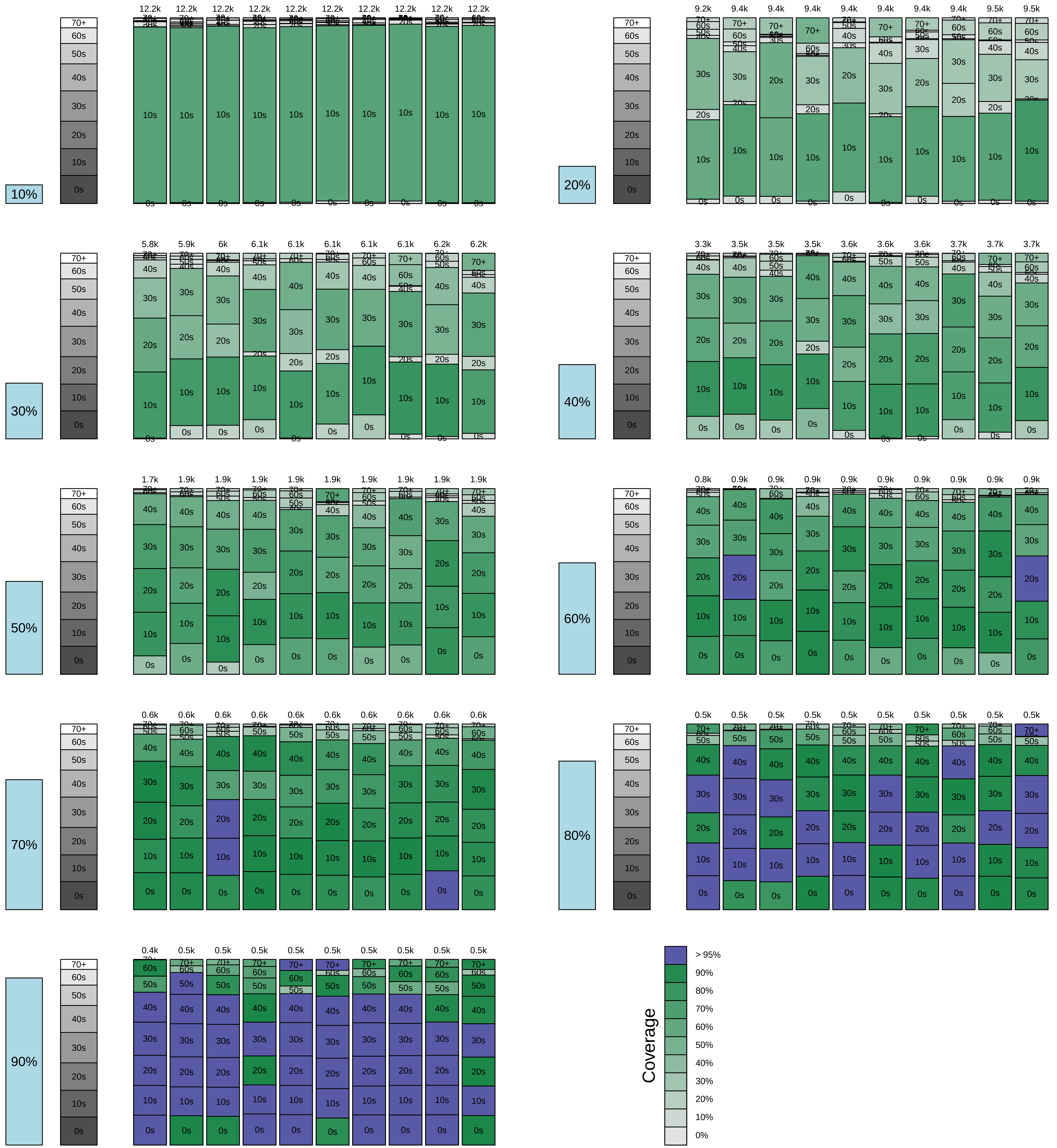

### 75% VN Population, 25% US Population

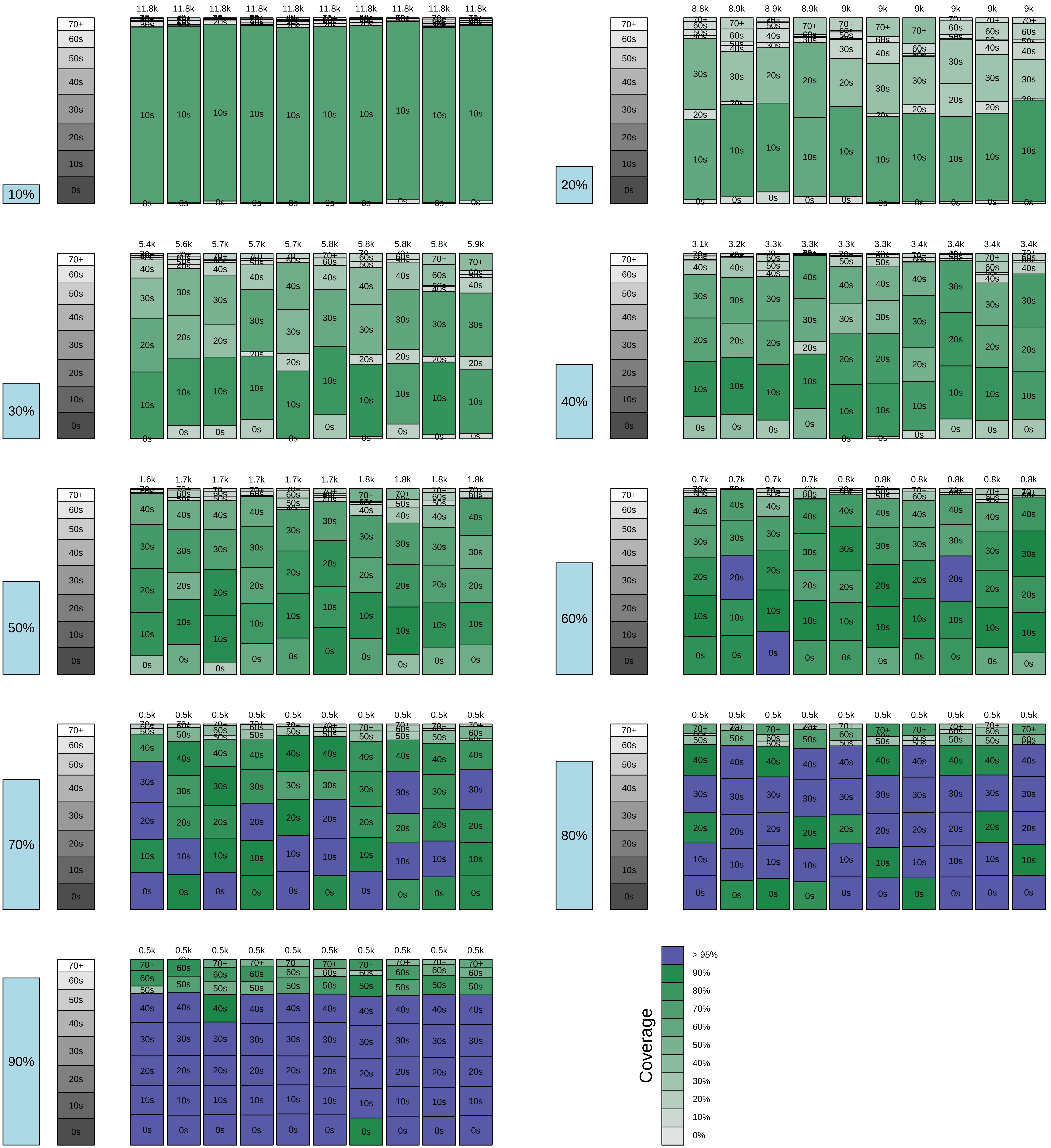

### 50% VN Population, 50% US Population

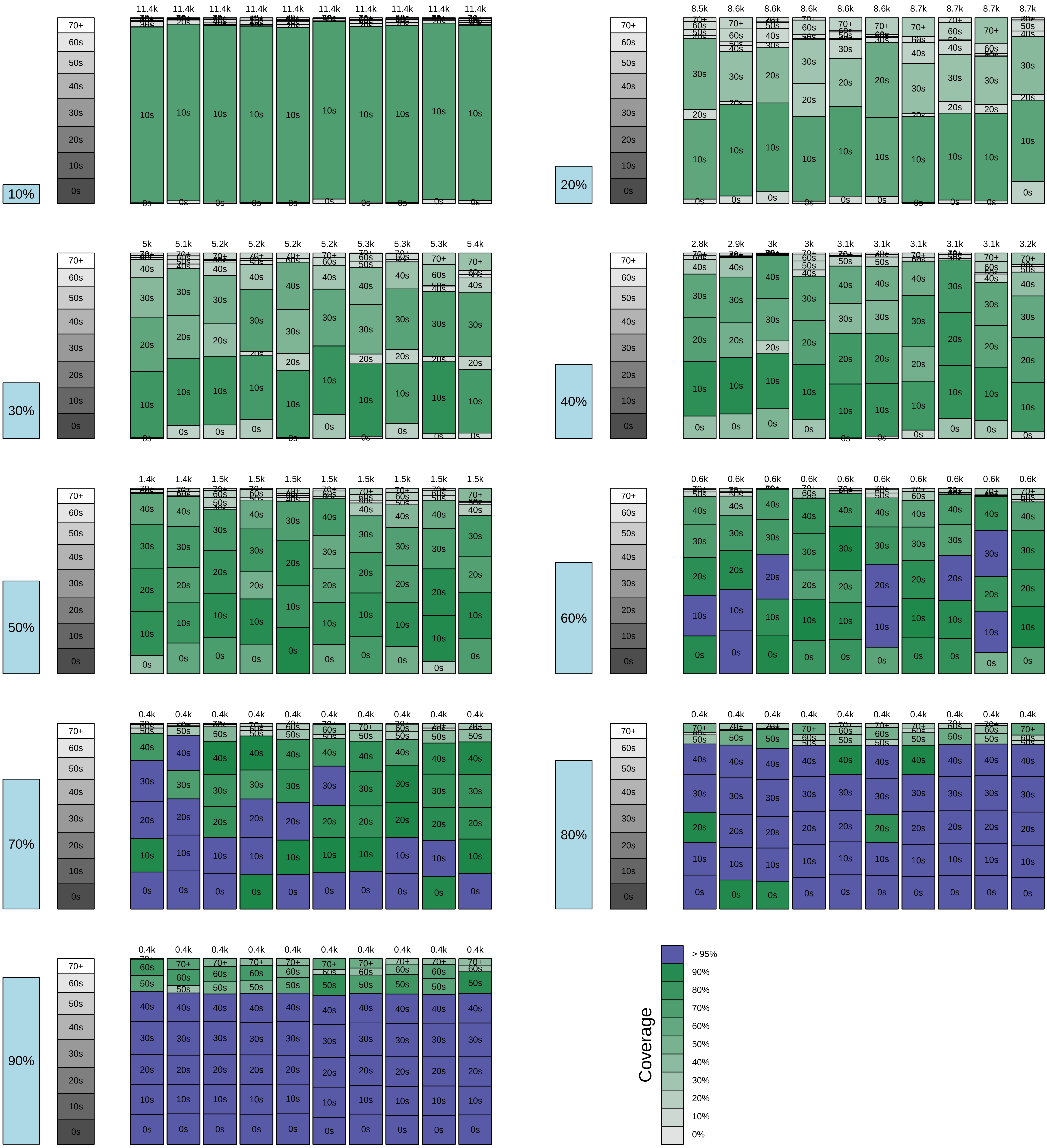

### 25% VN Population, 75% US Population

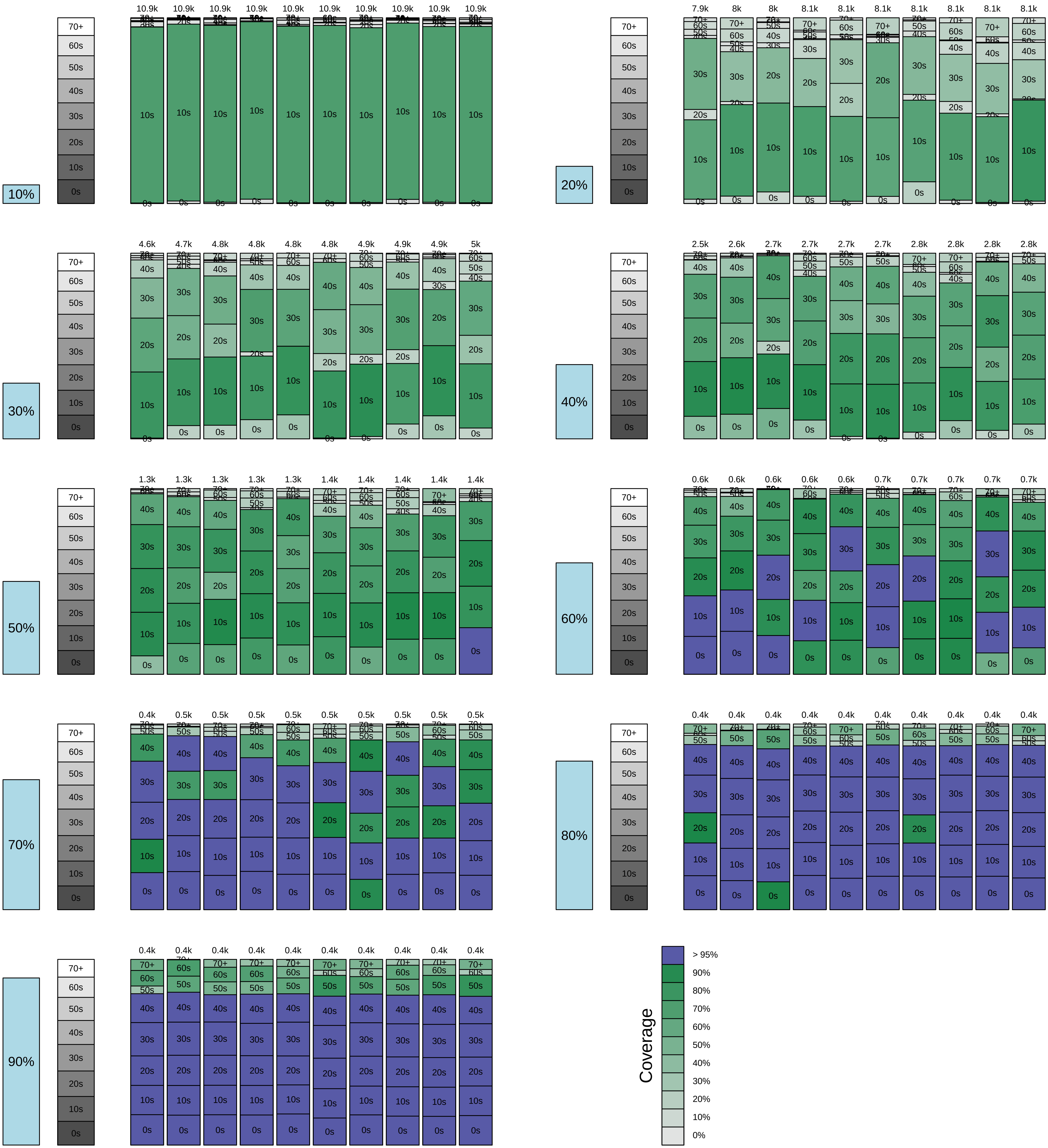

### 100% US Population

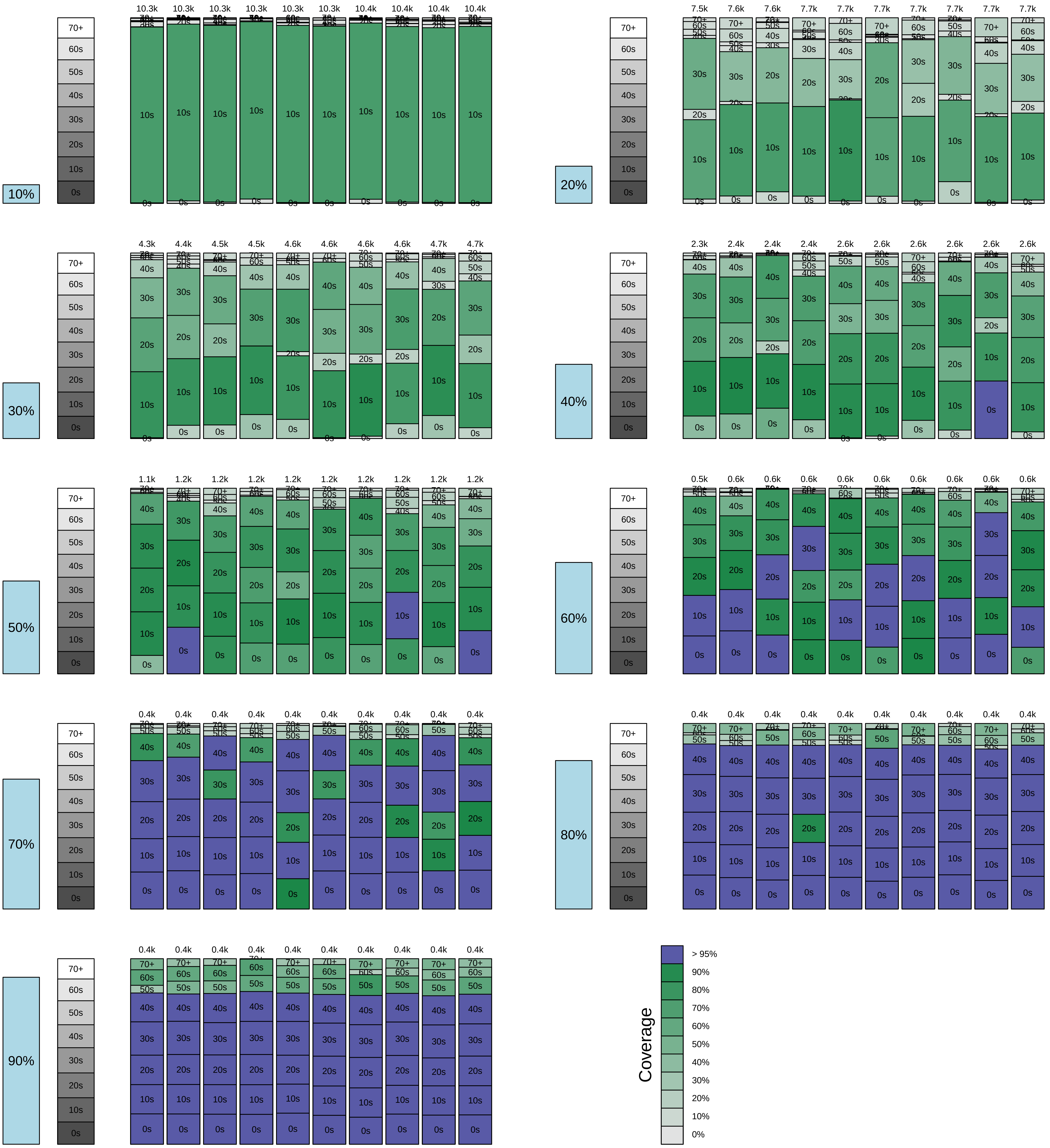
