## Supplemental Figure Captions for "Influenza vaccination allocation in tropical settings under constrained resources"

Figure S1. Model diagram used for analyses. This diagram represents the flow of compartments for each age group. Mixing across age groups is modeled based on previously estimated contact patterns. Compartments for Susceptible (S), Vaccinated (V), Infectious (I), second-stage infected ($I^2$), Hospitalized (H), Recovered (R), and Dead (D) are present. Compartment labels preceded by a lowercase v indicate Infectious, Hospitalized, or Recovered after being vaccinated. Subscript H1, H3, and B denote the (sub)type of infection. The circled number 4 over the Recovered stages indicates the presence of four Recovered compartments, with reinfection from the other (sub)types possible in any of these stages. Red arrows indicate vaccination, moving populations into the Vaccinated class. This diagram is repeated for all eight age classes.

Figure S2. Results from selecting optimal timing of vaccines. Each panel shows the average annual mortality at a given vaccine coverage when beginning vaccination at the specified month and administering vaccines over a span of one to four months. Blue lines indicate average mortality when vaccinating continuously over 12 months. Under each start month and duration, population coverage over the calendar year is equal.

Figure S3. Rankings of evaluated vaccine allocations. Allocations are ranked by average annual population mortality. Each page represents a different vaccine supply. Red dots represent an age-proportional vaccine allocation. Top panel: Average annual mortality for each vaccine allocation. Second through ninth panels: Vaccine coverages for each age group among the ranked vaccine allocations.

Figure S4. Optimal age-based vaccine allocations under different assumed vaccine effectiveness values. Mortality minimizing strategies based on vaccine supply if vaccine effectiveness against infection, hospitalization if infected, and death if hospitalized ranges between 30% and 70% are shown. Similar to Figure 4, blue bars indicate vaccine supply relative to the population, and relative proportions of the age groups in the general population of Vietnam are compared to the proportions of vaccines allocated to age groups. The ten strategies shown represent those associated with the lowest average annual mortality. The bottom-right panel shows the strategy at each vaccine supply associated with the lowest average annual mortality, scaled based on vaccine supply.

Figure S5. Shares of age groups comprising the set of ten mortality-minimizing vaccine allocations under different assumed vaccine effectiveness values across vaccine supplies. The average proportion of vaccines allocated to each age group among the vaccine allocations that lead to lowest average annual mortality are shown, by vaccine supply, for each assumed vaccine effectiveness to show how the most effective distributions of vaccines changes based on effectiveness.

Figure S6. Optimal age-based vaccine allocations under different assumed vaccine-induced immunity durations. Mortality minimizing strategies based on vaccine supply if immune duration ranges between 180 days and 360 days are shown. Similar to Figure 4, blue bars indicate vaccine supply relative to the population, and relative proportions of the age groups in the general population of Vietnam are compared to the proportions of vaccines allocated to age groups. The ten strategies shown represent those associated with the lowest average annual mortality. The bottom-right panel shows the strategy at each vaccine supply associated with the lowest average annual mortality, scaled based on vaccine supply.

Figure S7. Shares of age groups comprising the set of ten mortality-minimizing vaccine allocations under different assumed durations of vaccine-induced immunity across vaccine supplies. The average proportion of vaccines allocated to each age group among the vaccine allocations that lead to lowest average annual mortality are shown, by vaccine supply, for each assumed duration of immunity to show how the most effective distributions of vaccines changes based on immune duration.

Figure S8. Optimal age-based vaccine allocations under different age compositions in a population. Mortality minimizing strategies based on vaccine supply are shown based on age compositions of Vietnam, the United States, and intermediate values. Similar to Figure 4, blue bars indicate vaccine supply relative to the population, and relative proportions of the age groups in the general population of Vietnam are compared to the proportions of vaccines allocated to age groups. The ten strategies shown represent those associated with the lowest average annual mortality. The bottom-right panel shows the strategy at each vaccine supply associated with the lowest average annual mortality, scaled based on vaccine supply.

Figure S9. Shares of age groups comprising the set of ten mortality-minimizing vaccine allocations under different population age structures across vaccine supplies. The average proportion of vaccines allocated to each age group among the vaccine allocations that lead to lowest average annual mortality are shown, by age structure on a gradient between the age structures of Vietnam and the United States, for each assumed duration of immunity to show how the most effective distributions of vaccines changes based on the age demographics of the population.
